# The Variance-Stabilizing Transformation for the Poisson Rate Ratio: Closed-Form Confidence Intervals

**DOI:** 10.64898/2026.07.16.26358255

**Authors:** Siew-Phang Ng

## Abstract

The incidence rate ratio *R* is the standard measure for comparing event rates in clinical trials and epidemiology. In vaccine trials, the vaccine efficacy is VE = 1−*R*. When events are rare, the two arm counts are Poisson. The estimator of *R* is heteroskedastic: its sampling variance changes with the data. So no fixed-width interval covers correctly everywhere. The usual log-Wald interval is undefined at zero events and covers poorly at small counts. Early vaccine and drug-safety readouts fall in exactly this regime. We show that a single reparameterization collapses this bivariate problem to an effective one-parameter family with a quadratic variance function, whose variance-stabilizing transformation is 2 arcsinh 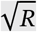. The reduction yields a closed-form confidence interval for *R*. Its two leading errors, a curvature bias and the variability of the estimated scale, each admit a closed-form correction with no tuning constants. In a Monte Carlo study of our seven arcsinh variants and five competitors, the +Curve+Stu variant covers within 0.002 of the nominal 0.95 for about 50 control and 5 treatment events. Its width is on par with the best competitor. It avoids the conservatism and zero-count breakdown of log-Wald and MOVER. For moderate counts, we recommend this interval; for sparser data, our Bar-Lev–Enis count-shift variant is more robust. The result is a ready-to-use, closed-form interval for the low-count regime. We illustrate it on early Covid-19 vaccine-efficacy readouts and provide reference implementations in R and Python.

## 1 Introduction

The ratio of two event rates is a core measure in clinical trials and epidemiology, arising whenever incident counts in a treatment arm and a control arm are compared. Beyond medicine, it appears in reliability engineering and the experimental sciences. In vaccine efficacy trials, the incidence rate ratio *R* is the primary parameter of interest, with vaccine efficacy VE = 1 − *R*. The Poisson rate-ratio is the natural framework for these trials ^1^, preferred over a binomial (risk-ratio) one. With many subjects, each at low event probability, the arm-specific case counts are Poisson (the law of rare events). Under staggered entry and variable follow-up, the estimand that matches the data is the incidence *rate* ratio, not a risk ratio tied to a fixed number at risk. Pfizer estimated its vaccine efficacy from the incidence rate ratio ^2^, Moderna from a Cox proportional-hazards model ^3^. Neither pivotal Covid-19 vaccine trial used a risk ratio.

### 1.1 The heteroskedasticity problem

While the Poisson rate ratio is the natural estimand, its estimator suffers from *heteroskedasticity*. Its sampling variance is not constant but changes with the data.

Throughout this subsection, to highlight the problem, we assume equal exposure times in the two arms. The unequal case reduces to this by the rescaling of Section 1.3. Let *K* ~ Pois(*μ*_1_) and *L* ~ Pois(*μ*_0_) be independent Poisson counts from the treatment and control arms, with means *μ*_1_ = *E*[*K*] and *μ*_0_ = *E*[*L*]. The estimand is the rate ratio *R* = *μ*_1_/*μ*_0_, estimated by 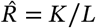.

The delta-method variance of 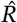 is estimated from the counts by

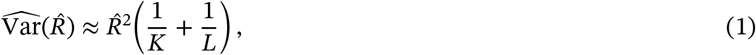

which varies with 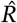. This dependence on the unknown *R* means no single fixed-width interval can achieve the stated coverage uniformly. A Wald-type interval calibrated at one *R* will be too wide or too narrow at another.

The standard workaround is the *log-Wald* interval: on the log scale the 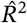 factor drops out, leaving 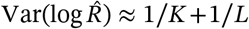, whence

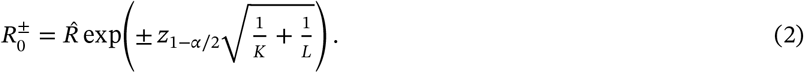

This first-order log-transform interval has several deficiencies: it is undefined when *K* = 0, systematically conservative in conditional coverage, and poorly approximated by the normal for small counts. Moreover, log is not the optimal transformation for the ratio. The stabilized variance 1/*K* + 1/*L* still depends on the data, especially when *K* is small.

### 1.2 Variance-stabilizing transformations

Since log is not the optimal transform, we step back and consider the *variance-stabilizing transformation* (VST). It is a monotone function *h* chosen so that 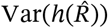 is approximately constant, regardless of the true *R*. A *pivot* is a function of the data and parameter whose distribution does not depend on nuisance parameters. The key advantage of the transformed scale is that a pivot can be formed on it directly from the standard normal distribution, yielding confidence intervals (CIs) with coverage that is uniform across the parameter space ^4^. The textbook example of a pivot is the studentized mean 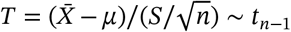, whose distribution is the same for every (*μ, σ*^2^). A confidence interval follows by inverting the probability statement *P*(|*T*| ≤ *t*_*n*−1, 0.975_) = 0.95 into a set of values for *μ*. The variance-stabilizing transformation plays the same role for the rate ratio, supplying an (approximate) standard-normal pivot from which the interval is read directly. The general construction, due to Bartlett ^5^, solves the differential equation 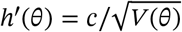 where *V*(*θ*) is the variance function. Informally, the transformation stretches the scale where the variance is large and compresses it where the variance is small, producing a uniform “ruler” for measuring deviations.

For the Poisson distribution itself, the classical VST ^4^ is 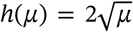, which stabilizes 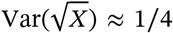 for *X* ~ Pois(*μ*). We shall see in Section 1.5 how to arrive at the VST of the *ratio* of two Poisson means.

### 1.3 Model and parameterization

Recall the counts *K* ~ Pois(*μ*_1_) and *L* ~ Pois(*μ*_0_). The counts are discrete, *K, L* ∈ {0, 1, 2, …}, while their means *μ*_*i*_ = *λ*_*i*_ *t*_*i*_ are continuous, fixed by the arm rates *λ*_*i*_ > 0 and the observed person-times *t*_*i*_ > 0. The estimand is the rate ratio

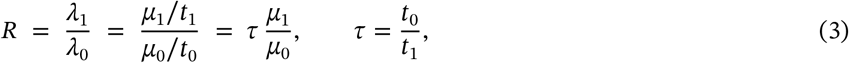

where *μ*_1_/*μ*_0_ is the ratio of expected counts, estimated by *K*/*L*, and *τ* is the known person-time ratio (under heterogeneous follow-up, *t*_*i*_ is the arm’s total person-time). The exposures enter only through this constant: any interval for the count ratio *μ*_1_/*μ*_0_ rescales by the positive factor *τ* to one for *R*, so its coverage transfers exactly. We therefore take equal exposures (*τ* = 1, *R* = *μ*_1_/*μ*_0_) without loss of generality, and recover the general case by multiplying the interval endpoints by *τ*. This treats *τ* as a fixed, known offset. Random or outcome-dependent person-time lies outside the model.

We introduce the *A-parameterization*:

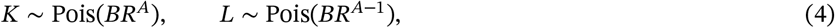

where *A* ∈ ℝ is a fixed splitting exponent and *B* > 0 is a nuisance (scale) parameter.^1^ Note that *E*[*K*]/*E*[*L*] = *R* for all *A*, so 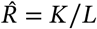 consistently estimates *R* regardless of the choice of *A*.

Setting *A* = 1 gives *K* ~ Pois(*BR*) and *L* ~ Pois(*B*), so that *L* is informative only about *B* and carries no information about *R*. This yields:

- The control count *L* ~ Pois(*B*) has a distribution free of *R*, so 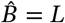 carries no information about *R*.
- Closed-form maximum-likelihood estimators 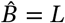 and 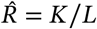.
- Conditional sufficiency: *K* ∣ *K*+*L* = *N* ~ Bin(*N, R*/(*R*+1)), which is *B*-free.

The control arm *L* now fixes the scale and nothing more. What is left is a one-parameter problem in *R*. The picture is an orbit reduced to its radius: two dimensions collapsing to one. We adopt *A* = 1 throughout the main development. The case for preferring it is made in Appendix A.

### 1.4 Variance of the rate ratio estimator

With the parameterization fixed at *A* = 1, the variance the transform must stabilize follows from the delta method applied to 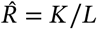 with *K* and *L* statistically independent:

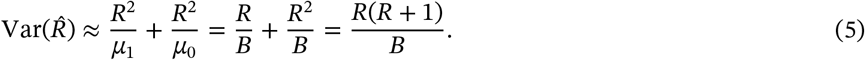

All coverage results below are conditional on the Poisson model: the variance 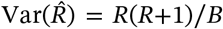 is the Poisson form. So under extra-Poisson variation (overdispersion) it understates the truth, and the intervals run narrow. Barker and Cadwell ^6^ also report this loss of coverage under negative-binomial data. A quasi-Poisson scale factor or the negative-binomial extension (§6.3) applies the same two-step refinement in principle, using a dispersion scale in place of the Poisson one. We do not evaluate coverage under overdispersion here.

### 1.5 The variance-stabilizing transformation for the rate ratio

The stabilizing recipe 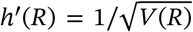 (Section 1.2, with *c* = 1) fixes the transform uniquely. Applied to the variance *V*(*R*) = *R*(*R* + 1)/*B* of (5), it gives 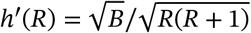, which integrates to

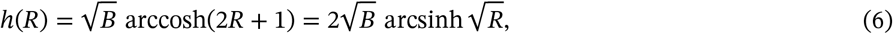

the second equality following from cosh(2*u*) = 2 sinh^2^ *u* + 1. On the *h* scale the delta-method variance is constant to leading order: 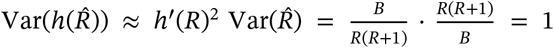, the *R*-dependence cancelling by construction. The unknown *B* enters only through the overall scale 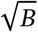. We therefore separate it from the *B*-free *kernel*

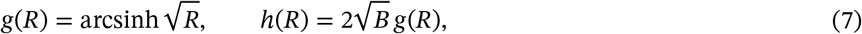

so that 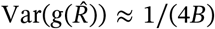. The formulas below are written in *g*, with the scale carried explicitly and 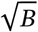 estimated by the observable 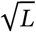. Since they use the arcsinh form (which complements the sinh^2^ inversion), we call this the *arcsinh CI* throughout.

### 1.6 Related work

The *A* = 1 parameterization reduces the problem to an effective single parameter. The counts are Poisson, but the estimator 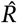 has variance *R*(*R* + 1)/*B*, quadratic in *R*. That is the negative binomial’s variance function. The reduction is unconditional. It keeps both counts and reads the scale off the *R*-free control arm, unlike the textbook conditioning on *N* = *K*+*L* that yields the binomial in *p* and discards the marginal. Our reparameterization and the elementary integration of Section 1.5 are self-contained.

The stabilizing arcsinh is familiar in the single-variate setting. It appears in Morris’s NEF-QVF catalog ^7^. Kulinskaya et al. ^8^ built variance-stabilizing confidence intervals for the whole class more than two decades later. Yu ^9^ applied the arcsinh to the negative-binomial mean. The closest methodological templates are both from the Kulinskaya–Staudte variance-stabilization program. The first is the arcsine VST for the binomial *difference* ^10^. The second, closer still in the transform itself, is an arcsinh VST of the Welch *t*-statistic for the standardized effect between two normal means ^11^, with the confidence interval obtained by inverting the stabilized pivot.

A separate strand applies the arcsinh to *binomial* effect ratios: Newcombe ^12^ shortens the logit interval for the odds ratio through an arcsinh map (the quarter-width image of the Wilson score interval). Fagerland and Newcombe ^13^ develop and evaluate it for the odds ratio and relative risk. These act as width transforms on the logit scale for the 2 × 2 table rather than as a variance-stabilizing transform of a rate ratio. None of these treats the Poisson rate *ratio*.

### 1.7 Contributions

The contributions are as follows.

1. The reparameterization that converts the *bivariate* Poisson rate-ratio problem into an effective *single-parameter*, quadratic-variance one: the move underlying the closed-form interval and its corrections (Sections 1.3–1.5).
2. An explicit closed-form CI for the Poisson rate ratio under the *A*-parameterization, including the studentization by 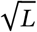 and interval inversion (Section 2).
3. A complete closed-form *refinement chain* for the arcsinh interval (Section 3; Tables 1 and 2). Each leading-order deficiency of the plain interval is traced to one mechanism and removed in closed form. The refinements fall into two camps, distinguished by where the correction enters: *pivot corrections* act on the standardized statistic the transformation produces (+Curve, +Curve+Stu, +Curve+Edge), and *count shifts* act on the counts before it (+Ansc, and +BLE, our rate-ratio equivalent of Anscombe’s 3/8 that removes the full leading-order bias). A one-sided fix handles the *K* = 0 boundary. The curvature correction and the count shift are alternative routes to the same leading bias; the studentization correction composes with either (e.g. +BLE+Stu).
4. A systematic Monte Carlo comparison of twelve CI methods across a (*R, B*) grid, establishing the operating characteristics (Section 4).
5. The argument that *A* = 1 is the preferred splitting exponent for rare events (*R* < 1) because of a clean *R*-free scale nuisance and the largest effective sample size in the sparse arm (Appendix A).
6. Illustration on early-readout Covid-19 vaccine efficacy data (Moderna and Pfizer-BioNTech) as a representative sparse-count example (Section 5).

**TABLE 1.**
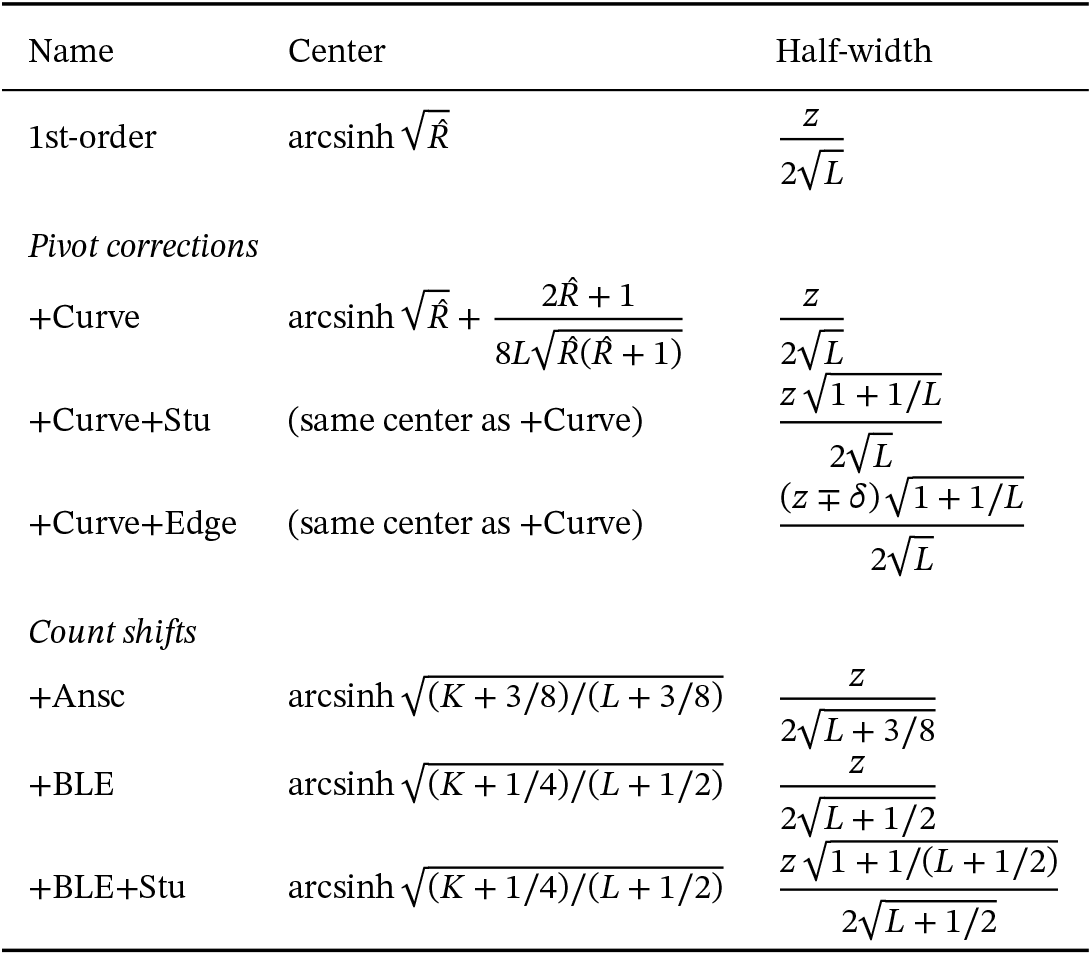
Refinement chain of the arcsinh interval family (methods (a)–(g)), grouped into pivot corrections and count shifts: the center and half-width of each interval on the 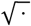 scale. Here *z* = *z*_1−*α*/2_, and 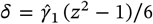 is the Cornish–Fisher skewness adjustment, defined with the estimated skewness 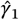 in Section 4 (Eq. (22)); it enters with *z* + *δ* on the lower bound and *z* − *δ* on the upper, hence the ∓.

**TABLE 2.** What each arcsinh refinement targets in the leading-order expansion. “✓” = mechanism explicitly addressed; “—” = not addressed. Methods (e) and (f) use count shifts that act on the estimator-bias and curvature terms *jointly*; the “jointly ✓” / “jointly partial” notation reflects that the shift affects both terms simultaneously rather than each in isolation.

| Method | Estimator bias | Curvature | Studentization | Skewness |
| --- | --- | --- | --- | --- |
| (a) 1st-order | — | — | — | — |
| (b) +Curve | — | ✓ | — | — |
| (c) +Curve+Stu | — | ✓ | ✓ | — |
| (d) +Curve+Edge | — | ✓ | ✓ | ✓* |
| (e) +Ansc | jointly partial<sup>†</sup> |  | — | — |
| (f) +BLE | jointly ✓ |  | — | — |
| (g) +BLE+Stu | jointly ✓ |  | ✓ | — |

\* Cornish–Fisher uses a plug-in estimate of $\gamma_1$ , which is noisy for $BR \lesssim 15$ and can degrade coverage in the sparse regime. <sup>†</sup> The shifts $(3/8, 3/8)$ act jointly on both terms; the residual leading bias is $(1+R)/[16B\sqrt{R(R+1)}]$ , strictly positive for all $R > 0$ : it never vanishes.

The construction is four steps.

1. **Reduce**. Setting *A* = 1 turns the two-count problem into a one-parameter one (Section 1.3).
2. **Stabilize**. The transform 2 arcsinh 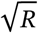 flattens the variance to a constant (Section 1.5).
3. **Read off**. Form a normal interval on the stabilized scale, then map it back to *R* (Section 2).
4. **Correct**. Recenter for the transform’s curvature, and widen the tails because the scale is estimated (Section 3).

The last step has exactly two parts. A symmetric interval can be wrong in only two ways: its center can be biased, or its half-width can miss the true scale.

## 2 The Arcsinh Confidence Interval

Section 1.5 established the stabilizing transform 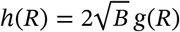, with 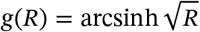. The interval then follows by working on the stabilized scale and inverting. The inversion is what makes the construction practical. For many variance-stabilizing transformations the inverse has no closed form, and the bounds must be located by root-finding. But *g*^−1^(*u*) = sinh^2^ *u* is monotone and trivially evaluated, so each bound comes from a single explicit formula.

### 2.1 The first-order confidence interval

The interval is read from a normal pivot on the stabilized scale. Define the (asymptotic) pivot:

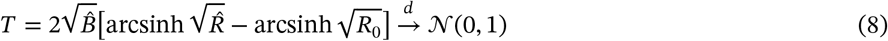

under *H*_0_ : *R* = *R*_0_, with 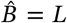. The convergence in (8) is asymptotic: the standard-normal pivot property is exact only in the limit *B* → ∞. At finite *B* the pivot carries *O*(1/*B*) corrections in both bias and variance, with an *R*-dependence that Section 3 addresses. The factor 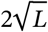 *studentizes* the pivot: it normalizes by an estimate of the standard deviation of the transformed estimator, analogous to dividing by 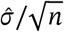 in the classical *t*-test. Inverting the pivot |*T*| ≤ *z*_1−*α*/2_ and using the monotonicity of arcsinh gives the closed-form CI:

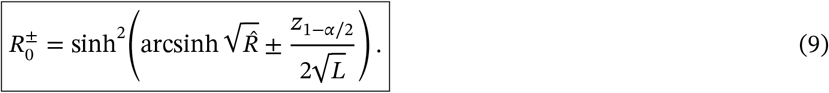

No numerical root-finding is required. When *K* = 0, 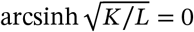, and both bounds collapse to the same point. The natural fix is to set the lower bound to 0, giving a one-sided 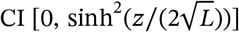. The corrected intervals of Section 3 inherit the same fix, each with its own half-width (Eq. (20)).

## 3 Second-Order Corrections

The first-order CI (9) is systematically liberal: conditional coverage falls to 0.906 at the extreme cell (*B, R*) = (25, 0.10), against a nominal 0.95 (Table 3). Most of that deficit is the *K* = 0 boundary treated in Section 4. A residual deficit remains at moderate expected counts (0.939 at *B* = 100, *R* = 0.06). We trace it to two independent mechanisms that degrade the interval in complementary ways, and correct each in closed form.

**TABLE 3.** Conditional coverage of twelve CI methods at nominal 0.95. Methods (a)–(g) are arcsinh variants (our refinement chain plus the two count-shift forms +Ansc and +BLE, and the orthogonal combination +BLE+Stu); (h)–(l) are competitors from the literature. Abbreviations: +C+S = +Curve+Stu, +C+E = +Curve+Edge, +A = +Ansc, +BLE = Bar-Lev–Enis-corrected arcsinh, +BLE+S = +BLE with studentization inflation, MOVER-R = MOVER-Rao, Log-W = Log-Wald, Grah = Graham score (Eq. 23), B-LR = Bartlett-corrected likelihood-ratio interval (Appendix B). Each entry is an exact Poisson-product sum (Section 4); the 10^7^-replicate Monte Carlo (seed 3125, SE ≈ 7 × 10^−5^) reproduces every entry to within Monte Carlo error.

| $B$ | $R$ | 1st-ord | +Curve | +C+S | +C+E* | +A | +BLE | +BLE+S | MOVER-R | MOVER-FT | Log-W | Grah | B-LR |
| --- | --- | --- | --- | --- | --- | --- | --- | --- | --- | --- | --- | --- | --- |
| 10 | 0.50 | 0.938 | 0.945 | 0.958 | 0.940 | 0.954 | 0.949 | 0.957 | 0.976 | 0.961 | 0.972 | 0.952 | 0.950 |
| 10 | 2.00 | 0.954 | 0.948 | 0.963 | 0.951 | 0.954 | 0.952 | 0.961 | 0.965 | 0.972 | 0.958 | 0.952 | 0.952 |
| 25 | 0.10 | 0.906 | 0.898 | 0.902 | 0.984 | 0.946 | 0.929 | 0.930 | 0.961 | 0.917 | 0.964 | 0.961 | 0.914 |
| 25 | 1.00 | 0.951 | 0.948 | 0.954 | 0.950 | 0.951 | 0.949 | 0.955 | 0.954 | 0.951 | 0.954 | 0.951 | 0.951 |
| 50 | 0.06 | 0.934 | 0.931 | 0.932 | 0.976 | 0.937 | 0.934 | 0.936 | 0.962 | 0.934 | 0.964 | 0.961 | 0.930 |
| 50 | 0.50 | 0.948 | 0.949 | 0.952 | 0.947 | 0.951 | 0.950 | 0.952 | 0.952 | 0.950 | 0.952 | 0.951 | 0.950 |
| 100 | 0.06 | 0.939 | 0.947 | 0.948 | 0.932 | 0.952 | 0.947 | 0.948 | 0.953 | 0.952 | 0.967 | 0.953 | 0.954 |
| 100 | 0.50 | 0.949 | 0.950 | 0.951 | 0.949 | 0.950 | 0.950 | 0.951 | 0.951 | 0.950 | 0.951 | 0.950 | 0.950 |
| 185 | 0.06 | 0.945 | 0.949 | 0.949 | 0.941 | 0.950 | 0.949 | 0.950 | 0.952 | 0.951 | 0.956 | 0.952 | 0.950 |
| 185 | 1.00 | 0.950 | 0.950 | 0.950 | 0.950 | 0.950 | 0.950 | 0.951 | 0.950 | 0.950 | 0.950 | 0.950 | 0.950 |
| 500 | 0.05 | 0.948 | 0.949 | 0.949 | 0.946 | 0.950 | 0.949 | 0.950 | 0.951 | 0.951 | 0.953 | 0.951 | 0.950 |
| 500 | 1.00 | 0.950 | 0.950 | 0.950 | 0.950 | 0.950 | 0.950 | 0.950 | 0.950 | 0.950 | 0.950 | 0.950 | 0.950 |

\* +Curve+Edge (like Log-Wald) conditions on $K \geq 1$ : its plug-in $\hat{\gamma}_1$ is undefined at $K = 0$ . At $B=25, R=0.10$ , where $P(K=0) \approx 0.08$ , its entry excludes the $K = 0$ boundary misses retained by the other arcsinh rows and is not comparable to them.

A symmetric interval 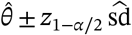 rests on two assumptions: that its center 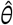 is unbiased for the parameter *θ*, and that its half-width matches the standard deviation. Bias breaks the first: the transformed estimate is shifted away from *θ*, so *θ* lies off-center in the interval, and coverage falls below the nominal 1−*α*. Random studentization breaks the second: calibrating the half-width with a single estimated scale leaves its tails too thin. The corrections below restore each assumption in turn.

### 3.1 Bias from curvature of the transformation

Take the first mechanism, the off-center bias. Recall the kernel 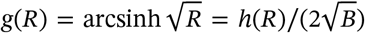 of (7), so the pivot is 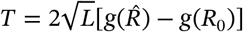. A second-order expansion of 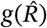 about the true value *R* has two *O*(1/*B*) contributions,

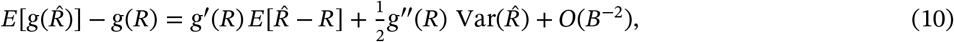

an estimator-bias term (the raw ratio is itself biased, 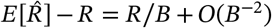)^2^ and a curvature term from the concavity of *g*. We remove the curvature term here (the +Curve correction); the estimator-bias term is left uncorrected. The count shift of Section 3.3 instead removes the full leading bias, both terms jointly. The accuracy of this expansion is controlled by the dimensionless curvature parameter

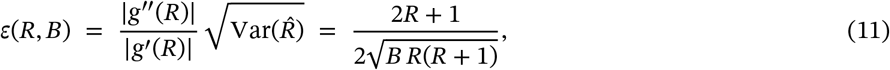

the second-order Taylor term being *ε*/2 times the first. For *R* of order unity, 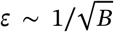, which is small whenever *B* is moderately large. For rare events (*R* ≪ 1), however, 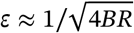, so the expansion requires *BR* ≫ 1, consistent with the *BR* ≥ 5 guideline stated in Section 4.

Computing *g*^′′^(*R*) = −(2*R* + 1)/(4[*R*(*R* + 1)]^3/2^) and substituting 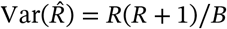, the curvature contribution is

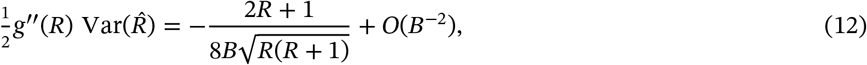

a downward shift of 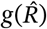. Adding the estimator-bias term 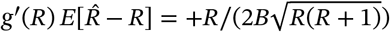 gives the net leading bias 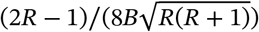, which changes sign at 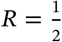. Section 4 works with this complete form.

The curvature-corrected estimator of *g*(*R*) subtracts the estimated curvature bias:

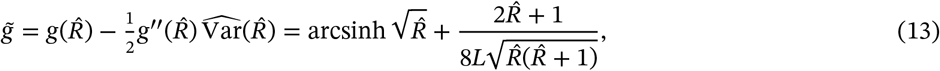

where 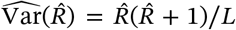 and the sign flip (subtraction of the negative *g*^′′^ term yields addition) is exact. This is the standard second-order (delta-method) bias correction, here applied on the variance-stabilizing arcsinh scale. On that scale the leading variance is data-independent, in contrast to the 1/*K* + 1/*L* of the log scale. The same standard correction on the log scale gives the ecological-meta-analysis form of Lajeunesse ^14^, ln(*K*/*L*) + 1/(2*K*) − 1/(2*L*) for Poisson data.

#### Remark 1

*Curvature is not the whole bias*

Both contributions are second order, but they are Jensen effects of *different* functions. The curvature term 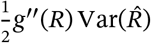 is the concavity of the transformation *g* acting on the spread of 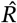. The estimator-bias term 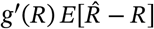 traces instead to the convexity of 1/*L*. Even with *K* ⊥ *L, E*[1/*L*] > 1/*E*[*L*], so 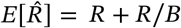. The +Curve correction estimates the first only; the Bar-Lev–Enis count shift (Section 3.3) cancels both jointly, which is why +BLE attains zero leading bias.

### 3.2 Studentization and the 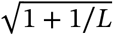 correction

The pivot (8) normalizes by the observable 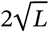 in place of the unknown 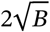. This substitution is *studentization*, the same step as replacing a known *σ* by 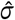 in the classical *t*-test. It leaves the pivot at unit variance to leading order. But it makes the sampling distribution *heavier-tailed*: a randomly estimated scale fattens the tails, exactly as it turns a *z* reference into a *t* reference. Here the single Poisson count *L* supplies the scale information. We widen the half-width by the factor

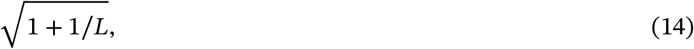

the tail-widening correction Student’s *t* makes for an estimated scale. It has no adjustable constant, and it holds coverage to nominal across the whole grid (Section 4). It acts on the tails rather than the variance: the studentized pivot retains unit variance to leading order.

### 3.3 The Bar-Lev–Enis count-shift

The two preceding refinements (+Curve and +Curve+Stu) shift the *pivot* additively to compensate for finite-sample bias and random studentization. An older tradition, going back to Anscombe ^4^, instead shifts the *counts* themselves before the VST is applied: *X* → *X* + *c* for some constant *c* chosen to optimize the finite-sample behavior of 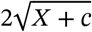. Anscombe’s optimal value is *c* = 3/8, which makes 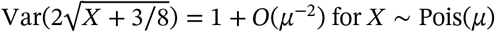.

For the rate-ratio problem the natural class of shifted estimators is

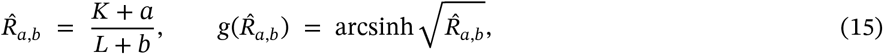

with two shift constants (*a, b*) that can in principle be tuned to target either the bias or the variance of the transformed pivot. The construction for the Poisson rate ratio, and the shifts derived below, are novel. They carry the Bar-Lev–Enis count-shift methodology ^15,7,16,17,18^ (single-variate in the classical literature) over to the bivariate ratio.

#### Choice of (*a, b*)

The leading 1/*B* bias of 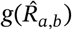, computed by a bivariate delta expansion in *ϵ*_*K*_ = *K* − *BR, ϵ*_*L*_ = *L* − *B*, is

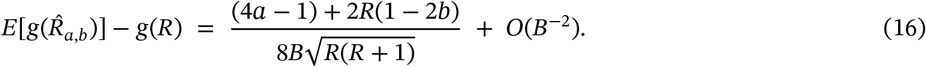

The numerator (4*a* − 1) + 2*R*(1 − 2*b*) is linear in *R*, so the bias vanishes *uniformly* in *R* only when both of its coefficients vanish: 4*a* − 1 = 0 from the *R*^0^ term and 1 − 2*b* = 0 from the *R*^1^ term. These are two conditions in the two free constants, with the unique solution

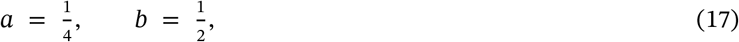

so that the shifted CI

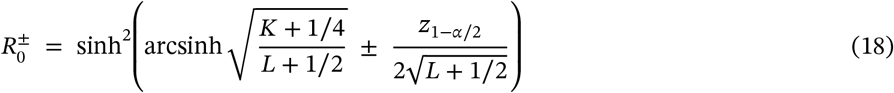

has zero leading bias uniformly in *R*. Thus (1/4, 1/2) is *derived* for the rate ratio by cancelling the leading bias (16), not transplanted from a single count. The shifts are uneven (unlike Anscombe’s symmetric 3/8) because the numerator and denominator contribute differently to the bias of the ratio. Notably, the same pair (*a, b*) = (1/4, 1/2) is the unique mean-matching solution for the univariate binomial arcsine VST in the general framework of Brown, Cai & DasGupta ^19^. The agreement reflects the conditional binomial structure *K* ∣ (*K* + *L*) ~ Bin(*K* + *L, R*/(*R* + 1)) at *A* = 1.

#### Comparison with +Ansc

The Anscombe-style shift (*a, b*) = (3/8, 3/8), which transplants the single-Poisson constant symmetrically to both counts, does *not* kill the leading bias of the bivariate transform: the residual is 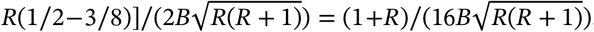, strictly positive for all *R* > 0. It never vanishes, so the symmetric transplant leaves an *O*(1/*B*) bias at every rate. Transplanting instead the single-Poisson *bias* shift symmetrically, (*a, b*) = (1/4, 1/4), removes the bias only in the rare-numerator limit *R* → 0. Its residual 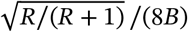 vanishes there and nowhere else. This is because the numerator wants a shift of 1/4 while the denominator wants 1/2. The shifts (1/4, 1/2) are the unique constants that cancel the leading ratio bias uniformly in *R*, generalizing the single-Poisson bias-canceling shift *a* = 1/4 (distinct from Anscombe’s variance-optimal 3/8) to the ratio’s two counts. Empirically (Section 4), the two count-shift variants give nearly identical coverage and width away from the *K* = 0 boundary, where the larger +Ansc numerator shift covers slightly better; both absorb the dominant finite-sample bias.

#### What the BLE shift does and does not achieve

The shift (17) cancels the leading *O*(1/*B*) bias identically in *R*, but not the next-order *O*(1/*B*^2^) residual. Both constants are already spent matching the two coefficients of the linear-in-*R* numerator of (16), and no freedom remains against the *R*-dependent residual. So (1/4, 1/2) is the parameter-count analog of Anscombe’s *one*-order cancellation, not the two-order cancellation that names the single-Poisson Bar-Lev–Enis optimum. Its explicit form is not needed here: the *O*(1/*B*^2^) residual enters through the finite-sample coverage of the simulation comparison, not as a term in the interval.

### 3.4 The fully corrected confidence interval

Combining the curvature correction (13) with the studentization inflation (14), the corrected +*Curve*+*Stu* CI is:

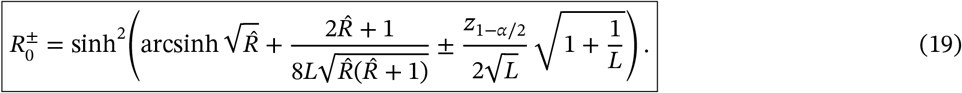

This interval is:

- *Closed-form*: no iteration or root-finding.
- *Parameter-free*: no tuning constants; the correction terms are derived.
- *Well-defined*: for *K* ≥ 1 and *L* ≥ 1 (with the *K* = 0 boundary handled by the one-sided fix 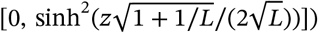.

It corrects the two second-order mechanisms it targets: transform curvature and random studentization. But it is not *O*(1/*B*)-unbiased. A residual estimator-bias term remains in its center.

#### Method hierarchy

The seven arcsinh refinements (methods (a)–(g), Table 1) fall into two camps, distinguished by where the correction enters. The *pivot corrections* (b)–(d) act on the standardized statistic the transformation produces. The *count shifts* (e)–(g) act on the counts before the transformation. The two camps are alternative routes to the leading bias, and the studentization step composes across them (+BLE+Stu). The five competitors (h)–(l) are introduced with the simulation study (Section 4).

The rows build in sequence, each adding the correction its name records (Table 1). One caveat is not visible in the formulas: simulation shows the noisy plug-in skewness estimate degrades coverage for small *BR*, making +Curve+Stu, not +Curve+Edge, the preferred closed-form method.

The complete recommended CI, combining the boundary fix with second-order corrections, is:

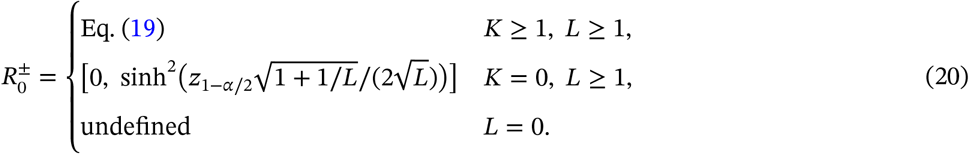

## 4 Monte Carlo Simulation Study

### 4.1 Design

Systematic comparisons of interval methods for the Poisson rate ratio are relatively few. Barker and Cadwell ^6^ compared eight in the rare-event regime, where the log-linear (log-Wald) interval was the narrowest of those maintaining coverage. The present comparison extends that study with the variance-stabilizing family and its second-order corrections.

Coverage probabilities are estimated via Monte Carlo simulation with the following design:

- *R*-grid: 25 points geometrically spaced from 0.01 to 5.0.
- *B*-grid: {10, 25, 50, 100, 185, 500}.
- 10^7^ independent replicates per grid point (1000 batches of 10,000, seed 3125, NumPy 2.4.6).
- Nominal level *α* = 0.05, *z*_0.975_ = 1.96.

Monte Carlo standard error per coverage entry is approximately 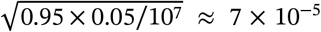. The coverage figures (Figures 2–4) are drawn from a companion scan at *B* ∈ {10, 25, 50, 100}, *R* geometrically spaced over [0.02, 1.0] (20 points), with 5 × 10^5^ replicates per point (MC SE ≈ 3 × 10^−4^).

**FIGURE 1.**
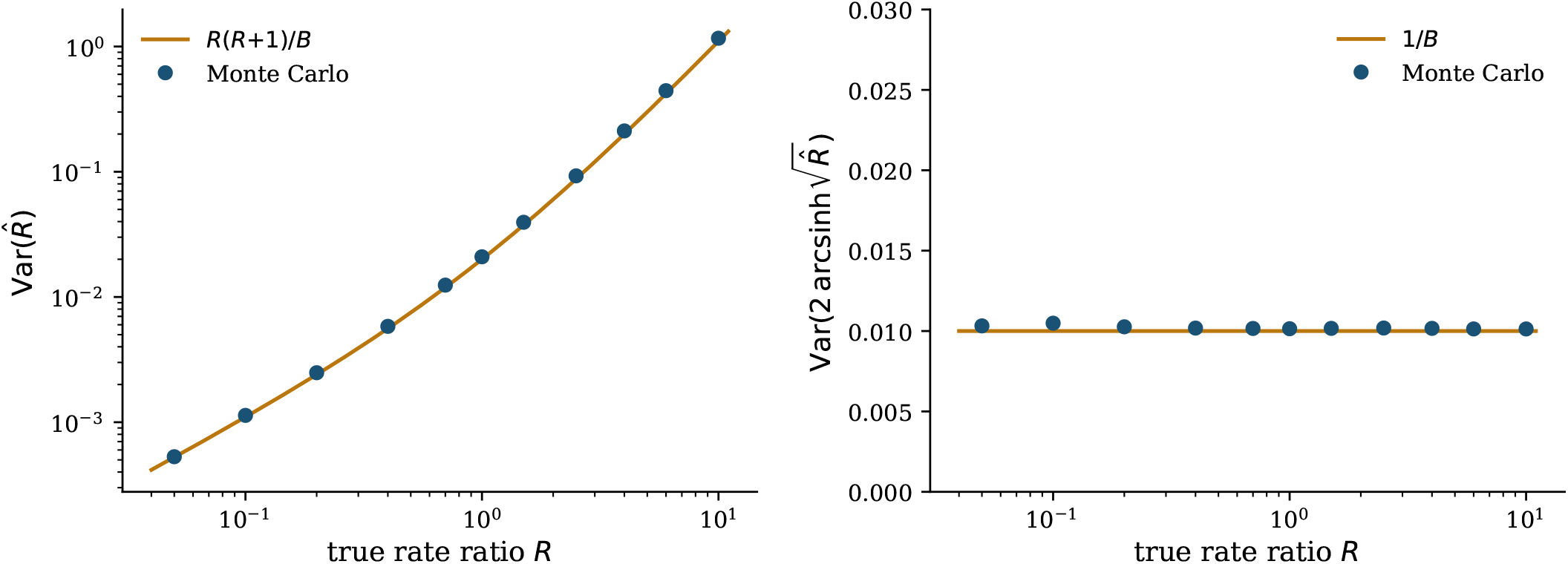
The variance-stabilization idea for the Poisson rate ratio, under the *A* = 1 parameterization *K* ~ Pois(*BR*), *L* ~ Pois(*B*) (here *B* = 100). *Left*: the delta-method variance of the raw estimator, 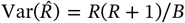, grows with *R*, so no fixed-width interval covers uniformly. *Right*: after the transform 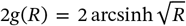, the variance is flat at ≈ 1/*B*, independent of *R*, a uniform scale on which a single normal quantile sets the interval half-width. Points are Monte Carlo estimates (4 × 10^5^ replicates per *R*); curves are the leading-order delta-method predictions.

**FIGURE 2.**
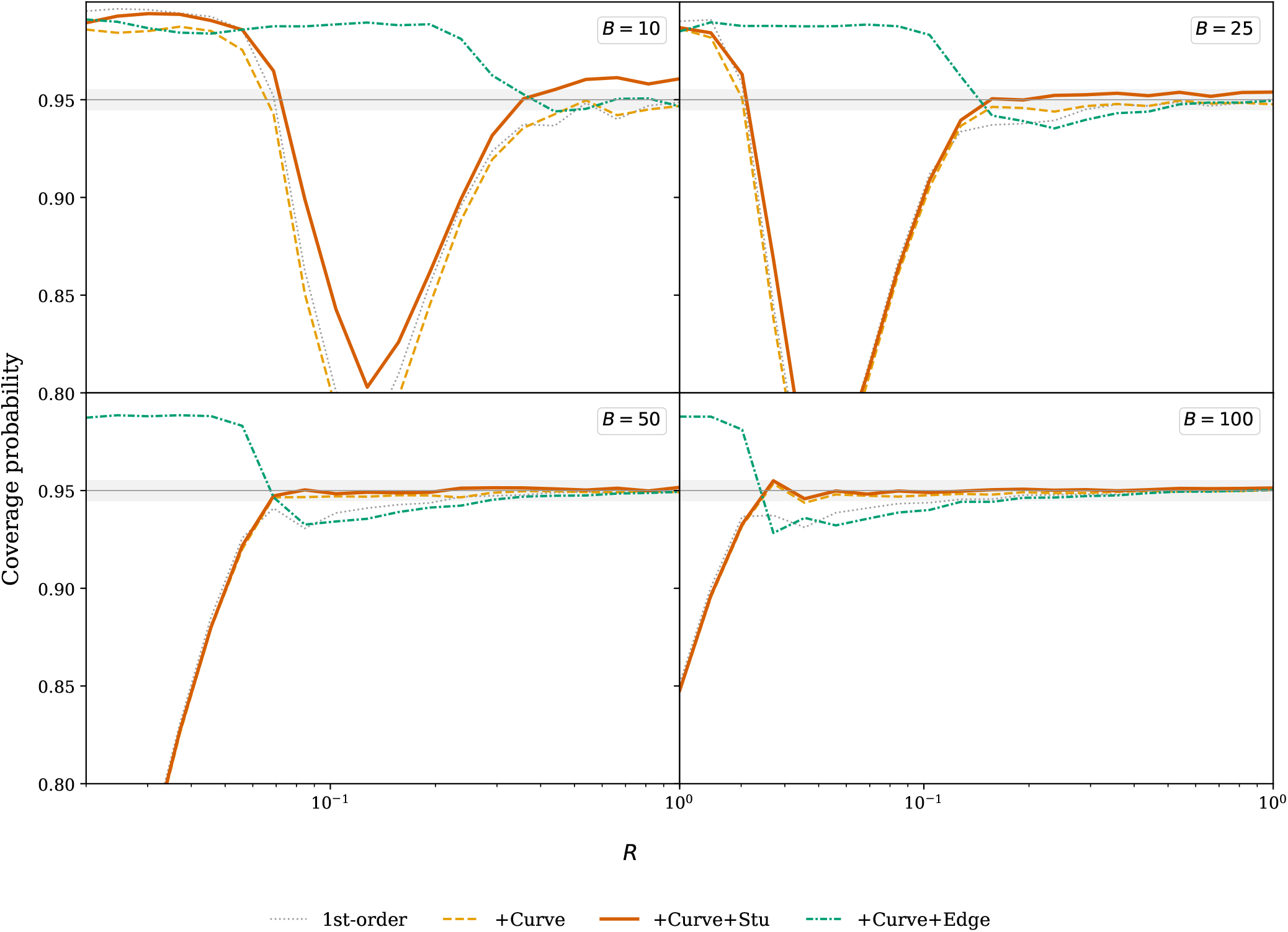
Conditional coverage of the arcsinh *pivot-correction* chain (*A* = 1) across (*B, R*): 1st-order (gray dotted), +Curve (orange dashed), +Curve+Stu (vermilion solid), and +Curve+Edge (green dash-dot-dot). These corrections act on the standardized pivot the transformation produces. The shaded band indicates 0.95 ± 0.005. Coverage improves through +Curve to +Curve+Stu, which achieves near-nominal coverage for *B* ≥ 50 and is the recommended closed-form interval; +Curve+Edge (Cornish–Fisher) is included only to show that it does *not* improve on +Curve+Stu: its plug-in skewness term degrades coverage at small *BR*.

**FIGURE 3.**
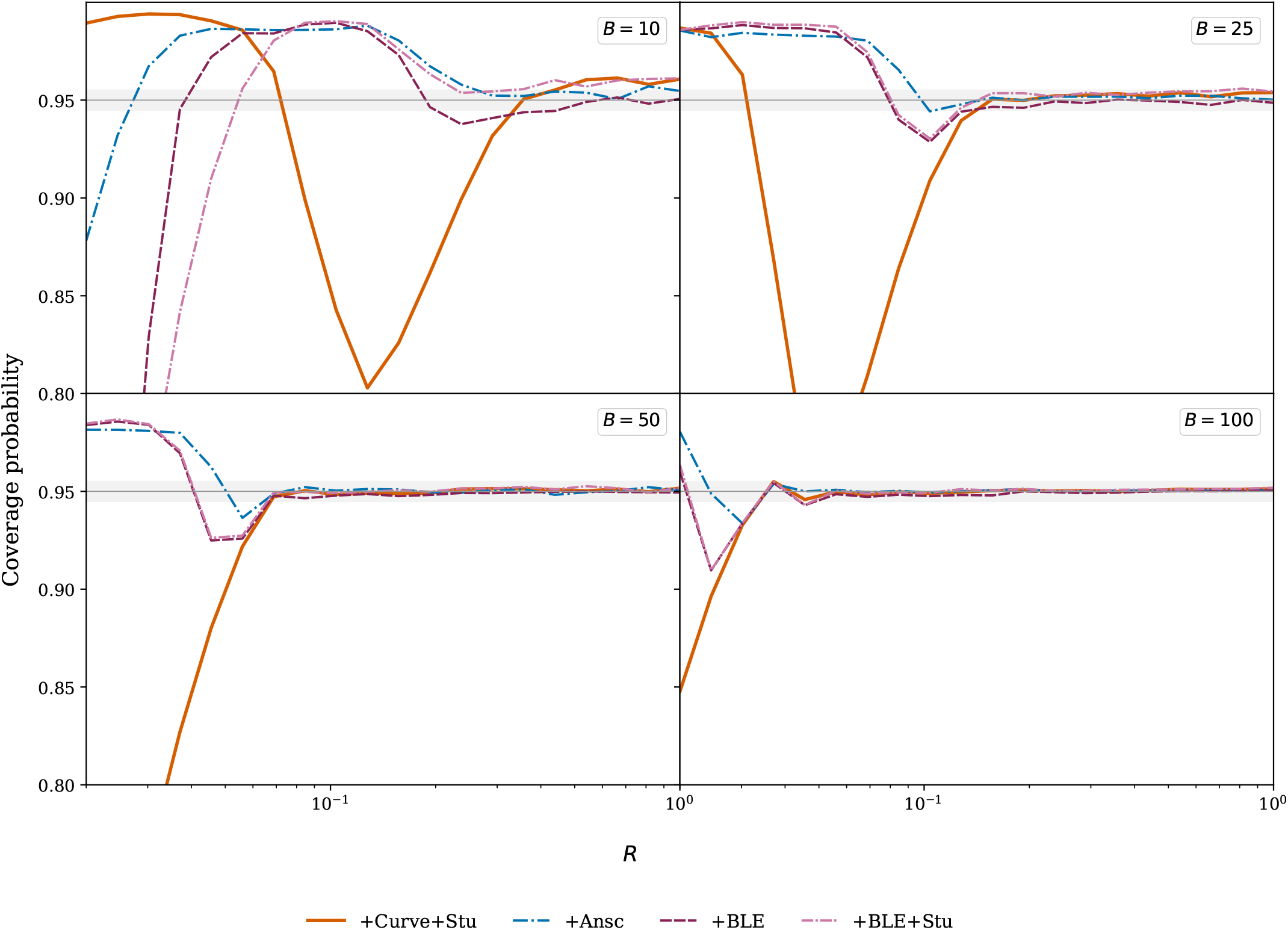
Conditional coverage of the arcsinh *count-shift* refinements (*A* = 1), against the recommended +Curve+Stu (vermilion solid) as reference: the Anscombe shift +Ansc (blue dash-dot), the Bar-Lev–Enis shift +BLE (wine long-dash), and the orthogonal combination +BLE+Stu (mauve long-dash-dot). These corrections act on the counts (*K, L*) before the transformation. The shaded band indicates 0.95 ± 0.005. The count shifts absorb the dominant finite-sample bias at the count level and stay near-nominal into the sparse regime (*B* ≤ 25) where the pivot corrections struggle; the +BLE shifts (1/4, 1/2) remove the full leading-order bias.

**FIGURE 4.**
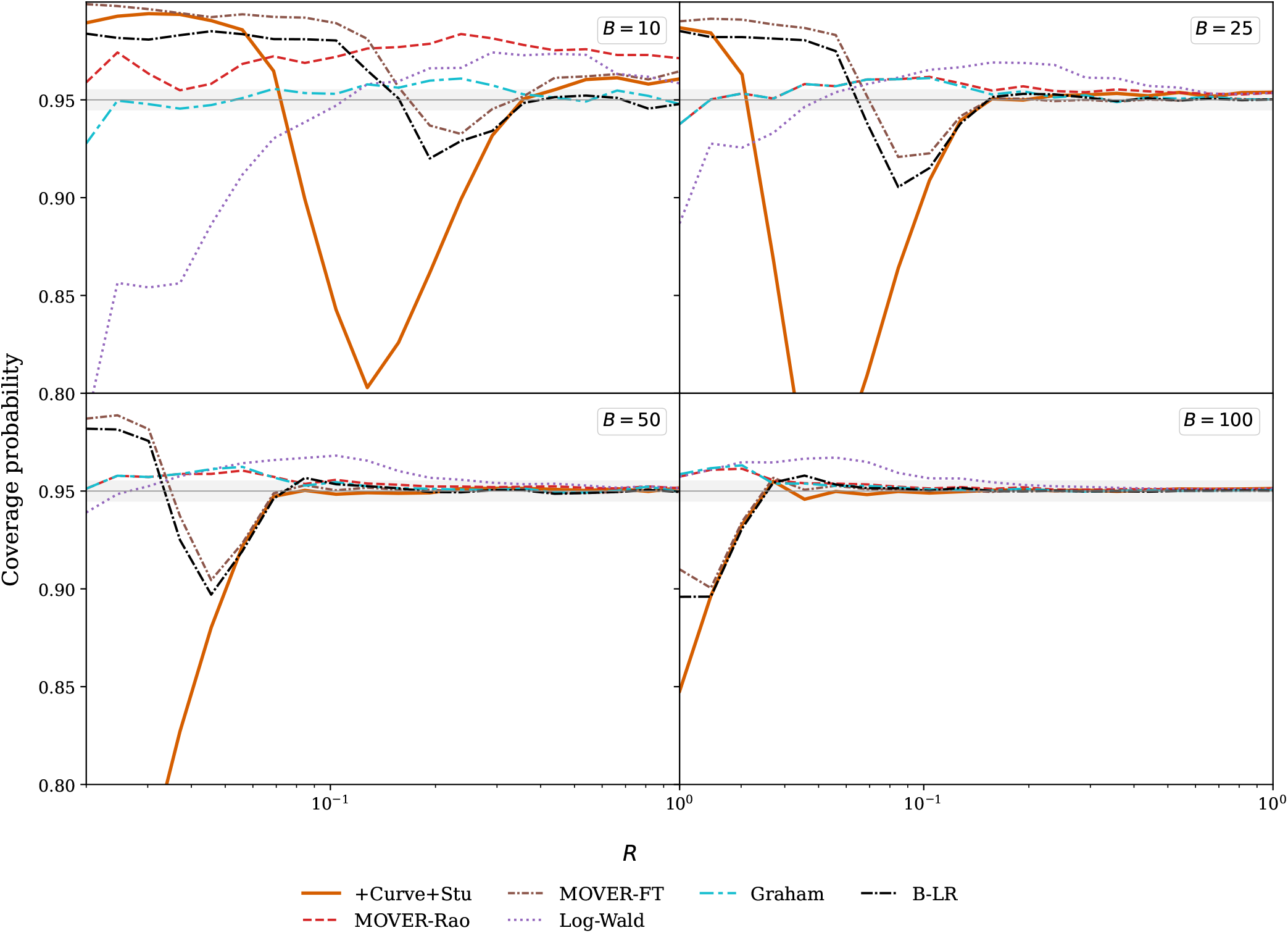
The recommended +Curve+Stu CI (vermilion solid) compared against the five literature competitors: MOVER-Rao (red dashed), MOVER-FT (brown dash-dot-dot), Log-Wald (purple dotted), the Graham score interval (cyan two-dash), and the Bartlett-corrected LR benchmark (B-LR, black dash-dot; Appendix B). The arcsinh count-shift forms are shown separately in Figure 3. For *B* ≥ 100, +Curve+Stu is near-nominal; MOVER-Rao is conservative and MOVER-FT can be liberal at small *BR*; Log-Wald is conservative and undefined at *K* = 0. The Graham score interval is near-nominal throughout and tracks MOVER-Rao closely for *B* ≥ 25, but, unlike Log-Wald, stays defined at *K* = 0. The B-LR benchmark is near-nominal for moderate counts but dips mildly liberal (to ≈ 0.90) in the sparsest cells, so even the root-finding second-order interval is not immune to the sparse-count regime.

Mean CI widths are *not* bounded. A ratio interval’s width grows without limit on the rare replicate where the denominator arm *L* is near zero. The width mean is therefore heavy-tailed, and its Monte Carlo estimate is seed-sensitive at small *B* (at *B* = 10 a 2 × 10^5^-replicate width ratio can wander a few percent between seeds). We therefore report all tables by *exact Poisson-product summation* over the count lattice (*K, L*) rather than by simulation: the coverage and per-tail tables (Tables 3 and 5), the width table (Table 4), and the *A* = 0 comparison (Table A1). Each Poisson factor decays super-exponentially, so truncating each arm at 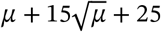 discards under 10^−12^ of the mass and returns the expectation to machine precision: a deterministic, seed-free value with no sampling error. The Monte Carlo design above generates the coverage figures and cross-checks the exact tables; it reproduces every coverage entry to within Monte Carlo error.

**TABLE 4.** Mean CI width *relative to +Curve+Stu*, at the same (*B, R*) points and for the same methods as Table 3 (> 1 wider, < 1 narrower; the +C+S column is 1.00 by construction). Widths are the mean over *defined* intervals (the conditioning of Table 3); for the *K*=0-undefined competitors at small *B* this averages over the defined subset only and therefore *understates* their width: at *B*=10, MOVER-Rao is undefined for 22% of samples and MOVER-FT for 13%, and log-Wald/+Curve+Edge reach 5–8% at the sparse *B*=25–50 points; the always-defined arcsinh and Graham forms carry no such bias. Each mean width is an exact Poisson-product expectation over (*K, L*), not a Monte Carlo estimate (Section 4): the width mean is heavy-tailed at small *B*, so a finite simulation is seed-sensitive there.

| $B$ | $R$ | 1st-ord | +Curve | +C+S | +C+E | +A | +BLE | +BLE+S | MOVER-R | MOVER-FT | Log-W | Grah | B-LR |
| --- | --- | --- | --- | --- | --- | --- | --- | --- | --- | --- | --- | --- | --- |
| 10 | 0.50 | 0.82 | 0.90 | 1.00 | 1.02 | 0.81 | 0.78 | 0.87 | 0.69 | 0.77 | 0.94 | 0.87 | 0.99 |
| 10 | 2.00 | 0.82 | 0.89 | 1.00 | 1.09 | 0.78 | 0.75 | 0.85 | 0.60 | 0.73 | 0.85 | 0.81 | 0.97 |
| 25 | 0.10 | 0.91 | 0.98 | 1.00 | 0.98 | 0.99 | 0.95 | 0.97 | 1.13 | 1.04 | 1.27 | 1.10 | 1.04 |
| 25 | 1.00 | 0.95 | 0.98 | 1.00 | 1.00 | 0.94 | 0.93 | 0.95 | 0.97 | 1.00 | 0.98 | 0.96 | 0.99 |
| 50 | 0.06 | 0.94 | 0.99 | 1.00 | 0.96 | 1.01 | 0.98 | 0.99 | 1.12 | 1.04 | 1.23 | 1.10 | 1.03 |
| 50 | 0.50 | 0.98 | 0.99 | 1.00 | 0.99 | 0.98 | 0.97 | 0.98 | 1.00 | 1.00 | 1.00 | 0.99 | 1.00 |
| 100 | 0.06 | 0.97 | 0.99 | 1.00 | 0.96 | 1.00 | 0.99 | 0.99 | 1.06 | 1.02 | 1.09 | 1.05 | 1.01 |
| 100 | 0.50 | 0.99 | 0.99 | 1.00 | 1.00 | 0.99 | 0.99 | 0.99 | 1.00 | 1.00 | 1.00 | 1.00 | 1.00 |
| 185 | 0.06 | 0.98 | 1.00 | 1.00 | 0.98 | 1.00 | 0.99 | 1.00 | 1.03 | 1.01 | 1.04 | 1.03 | 1.01 |
| 185 | 1.00 | 0.99 | 1.00 | 1.00 | 1.00 | 0.99 | 0.99 | 0.99 | 1.00 | 1.00 | 1.00 | 1.00 | 1.00 |
| 500 | 0.05 | 0.99 | 1.00 | 1.00 | 0.99 | 1.00 | 1.00 | 1.00 | 1.01 | 1.00 | 1.02 | 1.01 | 1.00 |
| 500 | 1.00 | 1.00 | 1.00 | 1.00 | 1.00 | 1.00 | 1.00 | 1.00 | 1.00 | 1.00 | 1.00 | 1.00 | 1.00 |

**TABLE 5.** Per-tail non-coverage at nominal 0.95: lower 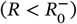 and upper 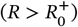 miss probabilities, with the asymmetry (upper − lower, from unrounded values). +Curve+Stu keeps near-symmetric tails for *R* ≥ 1 (the asymmetry shrinks with *B*); +BLE+Stu carries a persistent upper-tail excess that is flat in *R* and decays only with *B*: a mildly tight upper limit on *R*, hence on the VE lower bound. (Exact Poisson-product sums; the 10^7^-replicate Monte Carlo of `pertail_audit.py`, seed 3125, tail SE ≈ 5 × 10^−5^, reproduces every entry to within Monte Carlo error.)

| $B$ | $R$ | +Curve+Stu | | | +BLE+Stu | | |
| --- | --- | --- | --- | --- | --- | --- | --- |
|  |  | lower | upper | asym | lower | upper | asym |
| 25 | 1.00 | 0.020 | 0.025 | +0.005 | 0.012 | 0.033 | +0.020 |
| 100 | 0.50 | 0.023 | 0.026 | +0.003 | 0.020 | 0.029 | +0.010 |
| 100 | 1.00 | 0.024 | 0.026 | +0.002 | 0.019 | 0.029 | +0.010 |
| 100 | 2.00 | 0.024 | 0.025 | +0.001 | 0.019 | 0.030 | +0.010 |
| 185 | 0.06 | 0.021 | 0.029 | +0.008 | 0.020 | 0.030 | +0.010 |
| 500 | 1.00 | 0.025 | 0.025 | +0.001 | 0.023 | 0.027 | +0.004 |

### 4.2 Methods compared

We evaluate twelve CI methods: our seven refinements of the arcsinh family, (a)–(g) (the baseline (a), pivot corrections (b)–(d), and count shifts (e)–(g)), and five competitors, (h)–(l):

a. **First-order arcsinh**: Eq. (9) (uncorrected baseline).
b. **+Curve (curvature-corrected arcsinh)**: center shifted by 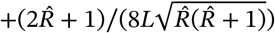
c. **+Curve+Stu (curvature- and studentization-corrected arcsinh)**: Eq. (19) (full second-order).
d. **+Curve+Edge (Cornish–Fisher-corrected arcsinh)**: the Cornish–Fisher expansion is a classical technique that adjusts normal quantiles to account for skewness (asymmetry) in the pivot distribution. Here it is applied on top of the fully corrected CI (19), using estimated skewness

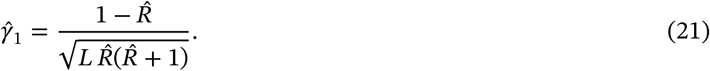

The Cornish–Fisher adjustment replaces the symmetric critical value *z* with asymmetric values

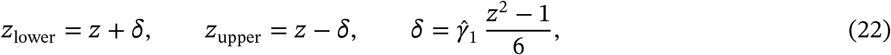

applied to the studentization-inflated half-width.
e. **+Ansc (Anscombe-corrected arcsinh)**: replace *K* → *K* + 3/8 and *L* → *L* + 3/8 in Eq. (9), giving 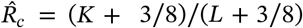 and half-width 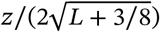. This is a continuity-corrected variant of the first-order arcsinh CI, transplanting Anscombe’s single-Poisson shift 3/8 symmetrically to both counts.
f. **+BLE (Bar-Lev–Enis-corrected arcsinh)**: replace *K* → *K* + 1/4 and *L* → *L* + 1/2 in Eq. (9), giving 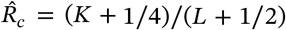 and half-width 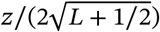. The shifts (1/4, 1/2) are the unique constants that kill the leading *O*(1/*B*) bias of 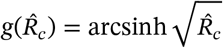 uniformly in *R* (Section 3.3); they are the rate-ratio equivalent of Anscombe’s 3/8 for the single Poisson, applied through the Bar-Lev–Enis ^7,16,17,18^ construction.
g. **+BLE+Stu (Bar-Lev–Enis with studentization inflation)**: the +BLE count shift combined with the studentization inflation 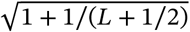 applied to the +BLE half-width. The two corrections target distinct mechanisms (count-level bias from the +BLE count shifts and pivot studentization from the studentization factor) and combine without double-counting; see also the decomposition argument in Section 6.
h. **MOVER-Rao score**: the method of variance estimates recovery with individual Rao score intervals, following Li et al. ^20^. It builds a score interval for each arm’s mean, then combines the two into an interval for the ratio.
i. **MOVER-Freeman-Tukey**: MOVER with individual Freeman–Tukey intervals ^20^ 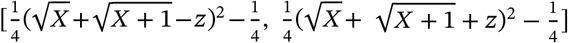.
j. **Log-Wald**: Eq. (2), the standard delta-method interval on the log scale.
k. **Graham score**: the non-iterative closed-form score interval of Graham et al. ^21^, obtained by inverting the score test for *R* with the nuisance rate profiled out (their eqs. 6–7). With *K* events in the numerator arm, *L* in the denominator arm, and 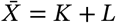, the bounds are

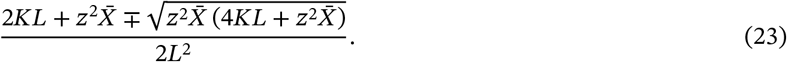

It is the nearest closed-form competitor to the present method and, unlike Log-Wald, is defined at *K* = 0 (returning [0, *z*^2^/*L*]); only *L* = 0 is degenerate.
l. **Bartlett-corrected LR (B-LR)**: the profile likelihood-ratio interval with the deviance Bartlett-corrected by the factor 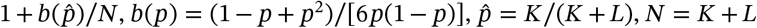. This is the textbook second-order-accurate benchmark; unlike the present method it requires root-finding. Its Bartlett coefficient is *R*-dependent and distinct from the 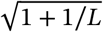 studentization factor; the construction and derivation are in Appendix B.

The exact conditional interval, Clopper–Pearson on *p* = *R*/(*R*+1) mapped through *R* = *p*/(1−*p*)^6^, is deliberately excluded from the twelve. It conditions on the total *N*, discarding the marginal, and over-covers by construction, so it is a conservative reference rather than a competitor. Its large-sample score form is the Graham interval (k) above.

#### What each refinement targets

Four independent mechanisms enter the pivot’s leading-order expansion at *O*(1/*B*), each derived above:

- the *estimator bias* 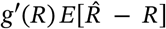 and the *curvature* 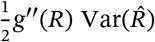, which combine to the net leading bias 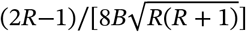 (Section 3.1);
- the *random studentization* tail correction 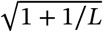 of Student-*t* type, with *L* in the role of the degrees of freedom (Section 3.2);
- the *pivot skewness* 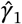 (Eq. (21)), which Cornish–Fisher (Eq. (22)) removes by adjusting the critical values.

Table 2 records which of the four each refinement addresses.

#### Expansion parameter

The Taylor expansion underlying methods (b)–(g) proceeds in the formal small parameter 1/*B*. Its leading bias is *O*(1/*B*), but the *R*-dependent prefactor diverges as *R* → 0, a first sign of the 1/(*BR*) control below. The studentized pivot keeps Var(*T*) ≈ 1 to leading order: the residual is a small *O*(1/*B*) tail effect, not a 1 + 1/*B* inflation. However, the practical accuracy regime is governed not by 1/*B* but by 1/(*BR*). The reason is that the relative standard error of 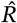 itself scales as 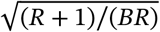. The Taylor expansion of any function of 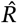 then has next-order terms with prefactors in inverse powers of *BR*, which become large for rare events (*R* ≪ 1). The two count-shift methods +Ansc and +BLE address this directly. Both leave the formal *O*(1/*B*) scaling intact, but +BLE’s shifts (1/4, 1/2) make the *O*(1/*B*) bias residual left by +Curve (the uncanceled estimator-bias term) identically zero for all *R*. For *R* → 0 this residual is small in absolute terms 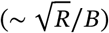 relative to the raw standard error 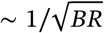 it is 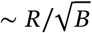 at fixed *R* and *O*(*R*^3/2^) at fixed *BR*. It is the residual that +BLE removes and +Curve does not, so +BLE holds an edge at small *BR*. The coverage *degradation* there, however, is driven by the expansion parameter 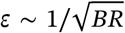 and the *K* = 0 boundary, not by this vanishing residual. +BLE’s joint cancellation of both mechanisms leaves no *O*(1/*B*) residual at all, so the first nonzero correction starts at *O*(1/*B*^2^). This is why +BLE outperforms +Curve in the sparse-event regime *BR* ≲ 5 even though both are nominally *O*(1/*B*)-correct methods.

Methods (h)–(j) operate outside this arcsinh framework: MOVER-Rao and MOVER-FT combine individual Poisson-mean CIs via Fieller-style recovery on the natural scale, and Log-Wald operates on the log scale rather than arcsinh. The categorization in Table 2 therefore does not extend to them. Their finite-sample behavior is governed by different approximations (Wilson-style score corrections, Freeman–Tukey stabilization, and the second derivative of log, respectively).

### 4.3 Results: conditional coverage

Table 3 reports *conditional* coverage (the fraction of defined intervals that contain the true *R*) at representative (*B, R*) points. We condition on *L* ≥ 1 (all methods); log-Wald and +Curve+Edge additionally require *K* ≥ 1 (the log scale is undefined at *K* = 0, and the plug-in skewness 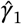 of Eq. (21) diverges at 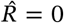). For the arcsinh methods other than +Curve+Edge, the only undefined case is *K* = 0, *L* = 0 (probability *e*^−*B*(*R*+1)^, negligible for *B* ≥ 10). For *K* = 0 with *L* ≥ 1, the one-sided boundary described below is used, so those samples are kept and the only excluded event is *L* = 0. For these arcsinh methods, unconditional and conditional coverage then differ by at most *P*(*L*=0) = *e*^−*B*^; +Curve+Edge, log-Wald, and the other *K* = 0-undefined competitors additionally drop *K* = 0, contributing *P*(*K*=0) = *e*^−*BR*^. Both bounds are < 10^−4^ for *B, BR* ≥ 10, with one exception: at *B* = 25, *R* = 0.10, *P*(*K*=0) = *e*^−2.5^ ≈ 0.08, so the +Curve+Edge entry there excludes the *K* = 0 boundary misses that the other arcsinh rows retain and is not directly comparable to them.

#### Behavior at *K* = 0

At the boundary *K* = 0 the interval is necessarily one-sided, and the methods differ in how far its upper limit reaches. The non-shift arcsinh variants take the one-sided boundary [0, sinh^2^(half-width)], each with its own half-width: 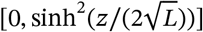 for the plain and +Curve forms (the +Curve center is singular at 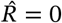, so the curvature shift is dropped at the boundary), and 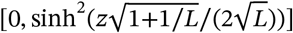 for +Curve+Stu, as in Eq. (20). All are ≈ [0, *z*^2^/(4*L*)], and the distinction does not affect the coverage in Table 3. At every tabulated (*B, R*), the true *R* exceeds all of these bounds except on a negligible set of *L*. Each rule therefore scores *K* = 0 as a miss. The count-shift forms keep a strictly positive center (*K*+*a*)/(*L*+*b*) and hence a higher upper bound. The Graham interval returns [0, *z*^2^/*L*]. The upper limits order as Graham > +Ansc > +BLE > the one-sided-fix arcsinh forms (the larger Anscombe numerator shift 3/8 > 1/4 lifting +Ansc above +BLE), so at fixed *L* they cover the true *R* out to successively larger *BR*. The Graham bound, for instance, covers up to *BR* ≈ *z*^2^ ≈ 3.8. This ordering explains the sparse-regime robustness of the count-shift and Graham methods. A wider upper bound at *K* = 0 covers the true *R* out to a larger *BR*, so these forms hold coverage further into the sparse regime than the plain arcsinh forms.

### 4.4 Width comparison and the coverage–width tradeoff

A confidence interval should be *valid* (coverage at least 1 − *α*; under-covering is *liberal*) and *precise* (as short as possible; needless over-coverage is *conservative*). We screen methods for validity first (coverage ≥ 0.945, no more than 0.005 below nominal), then rank the survivors by width.

Figures 5 and 6 and Table 4 give mean widths relative to +Curve+Stu; a wider-*B* scan (to *B* = 1000) confirms the pattern. The differences appear only at small expected counts, and the two families converge along different axes. The un-inflated count-shift forms +Ansc and +BLE are narrower than +Curve+Stu at moderate *R* and run at parity in the sparse small-*R* cells. The gap is the studentization inflation 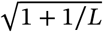 they omit: it tracks 1/*L*, and it is the price +Curve+Stu pays for holding coverage near nominal. The literature competitors are instead *wider* in the sparse small-*R* regime the application occupies, reaching 1.1–1.3× the +Curve+Stu width at the sparse points of Table 4. Their excess shrinks with the expected counts rather than with *B*, settling to within a percent or two once *BR* ≳ 25. The Bartlett-corrected LR benchmark is the tightest of the competitors.

**FIGURE 5.**
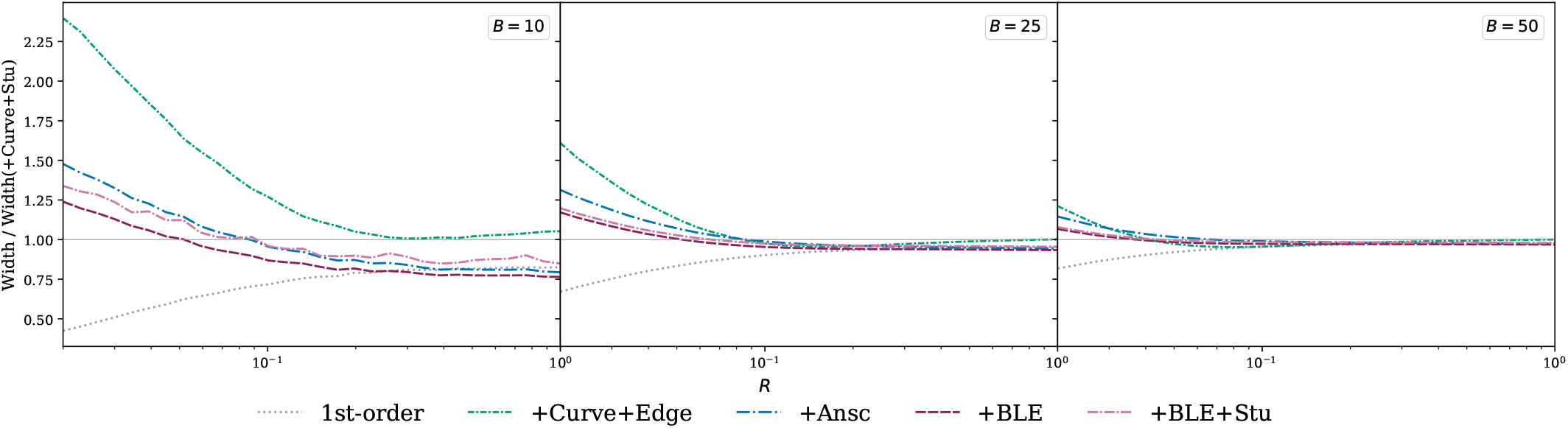
Relative CI width of the arcsinh *family*, normalized by the +Curve+Stu width (the gray line at 1.0), for small background rates (*B* ≤ 50): 1st-order (gray dotted), +Curve+Edge (green dash-dot-dot), +Ansc (blue dash-dot), +BLE (wine long-dash), and +BLE+Stu (mauve long-dash-dot). (+Curve is omitted for legibility: its width lies between 1st-order’s and +Curve+Stu’s, Table 4.) The +Ansc and +BLE count-shift for ms are the narrowest, staying competitive even in the sparse-event regime where the higher-order corrections break down; the studentization inflation 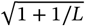 is the small price +Curve+Stu pays for holding coverage.

**FIGURE 6.**
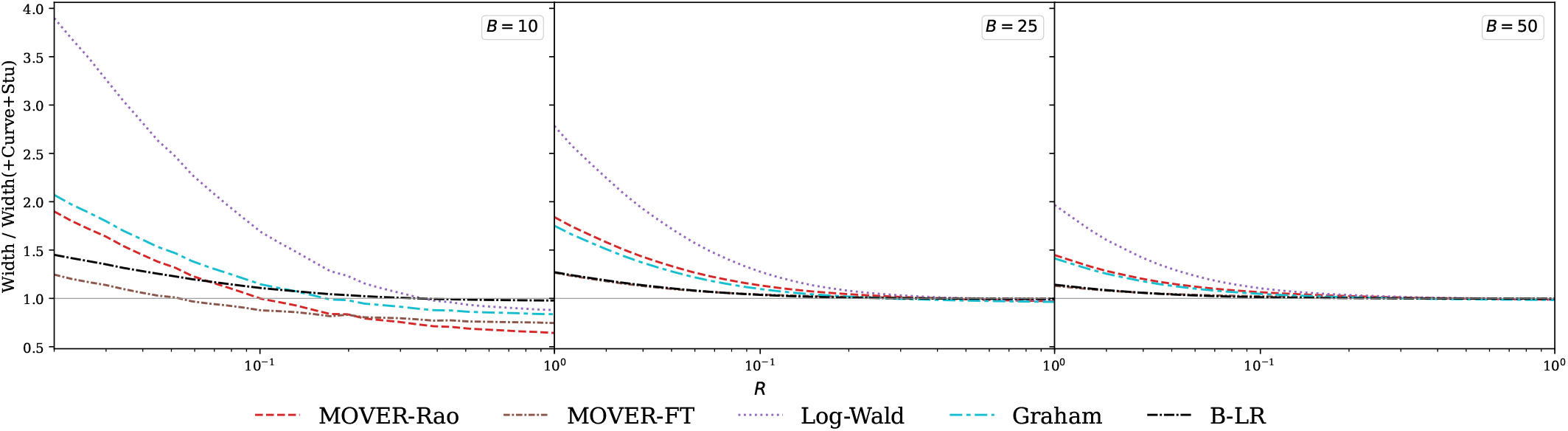
Relative CI width of the five literature competitors, normalized by the +Curve+Stu width (the gray line at 1.0), for small background rates (*B* ≤ 50): MOVER-Rao (red dashed), MOVER-FT (brown dash-dot-dot), Log-Wald (purple dotted), Graham (cyan two-dash), and the Bartlett-corrected LR benchmark (B-LR, black dash-dot). The competitors are wider than +Curve+Stu in the sparse small-*R* regime the application occupies, their excess shrinking with the expected counts rather than with *B*; B-LR is the tightest, running only slightly wider and converging as counts grow. Note the wider *y*-range than the arcsinh family of Figure 5.

#### Coverage–width tradeoff

In the intended regime (*B* ≥ 50, *BR* ≥ 5) the width ordering is stable: the un-inflated count-shift forms are marginally narrowest, +Curve+Stu sits a few percent above them, and the literature competitors are widest (most so at small expected counts). The 1st-order arcsinh is narrower still, but its liberal coverage (< 0.945) disqualifies it. +Curve+Stu is recommended in this regime not because it is the narrowest (it is not) but because its width is competitive while its principled studentization holds coverage closest to nominal in closed form. In the sparse regime (*B* < 50 or *BR* < 5) the count-shift forms +BLE and +Ansc are both narrower and better-covering, and are the recommended choice there (Section 4.7). This regime split also keeps the analysis comfortably outside the small-counts caution of Warton ^22^, whose theorem shows that variance-stabilization becomes impossible as the expected count approaches zero.

### 4.5 Key observations

Reading Table 3 method by method (Figures 2–4 plot conditional coverage across (*B, R*) from the companion Monte Carlo scan of the Design subsection):

1. The **1st-order arcsinh** CI is liberal where counts are sparse (0.906 at (*B, R*) = (25, 0.10)) and near-nominal elsewhere. The deficit vanishes as *B* → ∞. The **+Curve correction** removes 50–80% of that deficit for *B* ≥ 100, overcorrecting slightly at small *BR*.
2. The **+Curve+Stu correction** achieves within 0.002 of nominal for *B* ≥ 50 and *BR* ≥ 5. Below that regime its coverage is *non-monotonic* rather than a smooth decay. As *BR* falls, it dips to a trough, then climbs back *above* nominal (0.98–0.99) for *BR* ≲ 0.5. At the trough, the exact conditional coverage is 0.82 at *BR* = 1 for *B* = 25 and 0.73 for *B* = 185, bottoming near 0.75 and 0.70 at *BR* ≈ 1.25. The trough comes from the *K* = 0 boundary. The one-sided fix [0, sinh^2^(⋅)] ≈ [0, *z*^2^/(4*L*)] is a guaranteed miss when 1 ≲ *BR* ≲ 5, since the true *R* exceeds that upper bound. It always covers once *R* is small enough. The breakdown of the curvature-bias term compounds it. An exact Poisson-summation sweep across the boundary confirms this two-regime behavior.
3. **+Curve+Edge** does not improve on +Curve+Stu. Its plug-in skewness 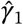 is too noisy for *BR* ≲ 15.
4. **+Ansc** and **+BLE** are nearly indistinguishable except where *K* = 0 carries mass: at *B* = 25, *R* = 0.10, the larger +Ansc numerator shift keeps a higher one-sided upper bound at *K* = 0, and +Ansc covers 0.946 where +BLE covers 0.929 (the ordering of the Behavior-at-*K* = 0 paragraph). Both are robust, staying within 0.021 of nominal even at that cell, where the higher-order corrections struggle. The cost is slight conservatism for moderate *B*. Both absorb the dominant finite-sample bias at the count level, before the VST is applied (+BLE by the bias-killing (1/4, 1/2) shift of Section 3.3).
5. **+BLE+Stu** combines the +BLE count shifts with the studentization tail inflation 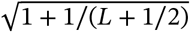 on the shifted scale. It is within 0.001 of +BLE for *B* ≥ 100 and slightly more conservative for *B* ≤ 25. Its per-tail asymmetry (Table 5) is taken up next.
6. Among the five literature competitors, **MOVER-Rao** and **Log-Wald** are conservative and never liberal, **MOVER-FT** turns liberal in sparse small-*R* cells, and **Graham score** is the best-behaved: near-nominal across the grid (0.950– 0.961) and, like the arcsinh forms, defined at *K* = 0. Two failure modes separate them from the present method. MOVER’s Fieller combination is undefined for a growing fraction of samples as *B* falls (~ 22% at *B* = 10, ~ 87% at *B* = 5), and Log-Wald is undefined at *K* = 0.

The Graham score interval is the natural closed-form benchmark, and across the recommended regime it matches the corrected arcsinh interval closely. The two reach that interval by different routes. Graham inverts the score test in a single step, standardizing by the expected information. The interval is calibrated by construction and stays defined at *K* = 0, though the finite-sample error it absorbs stays implicit in the algebra. The arcsinh construction instead names that error (a curvature bias and the cost of estimating the scale) as separate closed-form terms on a scale where the variance is constant, and the same construction produces the count-shift forms used when counts are sparse. The contribution is this decomposition and the family it generates, not an improvement in coverage over Graham.

### 4.6 Per-tail asymmetry

A two-sided interval can cover at the nominal rate while splitting its 5% miss unevenly between the two tails. That matters here because the regulatory decision rests on one tail only: the VE lower bound, equivalently the upper limit 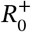 on the rate ratio. An interval with too tight an 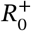 reports an optimistic VE lower bound and can clear the 30% threshold it should not, even when its two-sided coverage is nominal.

Table 5 splits the two-sided non-coverage into its lower 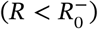 and upper 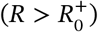 tails. +Curve+Stu and +BLE+Stu reach comparable two-sided coverage by different routes (per-method detail in the caption): the symmetric tails of +Curve+Stu are why it is preferred for the one-sided policy bound of Section 5.

### 4.7 Practitioner guidance

Based on the simulation results, we recommend:

- For *B* ≥ 50 and *BR* ≥ 5: use the fully corrected arcsinh CI (Eq. 19). It achieves coverage within 0.002 of nominal with a simple closed-form expression.
- For *B* < 50 or *BR* < 5: use +BLE+Stu (method (g)), +BLE (method (f), Section 3.3), the +Ansc arcsinh (method (e)), or MOVER-Rao score (method (h)). All four are more robust in the sparse-event regime, though at the cost of conservatism. +BLE and +Ansc give nearly identical intervals. +BLE has the cleaner derivation (bias-killing under the Bar-Lev–Enis ^17,18^ construction) while +Ansc uses the better-known Anscombe shift. +BLE+Stu adds the studentization factor on the +BLE-shifted scale and is the natural default when one wants the bias-killing property and the random-studentization correction simultaneously: it is the count-shift counterpart of +Curve+Stu.
- Avoid log-Wald for small event counts (*BR* < 5): it is undefined at *K* = 0 and has inflated conservatism.

The thresholds *B* ≥ 50 and *BR* ≥ 5 are on the expected counts (at least 50 control and 5 treatment events) and are invariant to the exposure times, since the law of (*K, L*) depends on them only through the means (Section 1.3).

## 5 Application: Covid-19 Vaccine Efficacy

We illustrate the methods on the two Covid-19 vaccine trials, Moderna ^3^ and Pfizer-BioNTech ^2^, taking equal exposure times for simplicity (the unequal case reduces to this by the rescaling of Section 1.3). These are the primary efficacy analyses that supported authorization. Their interest here is that the vaccine arm is intrinsically sparse: a vaccine that works produces few breakthrough cases, so the numerator count stays small (8–11 here) even though the trial has accrued its full complement of events. These readouts sit near the sparse *BR* ≈ 5–10 boundary that Section 4 maps: below it the methods separate, above it they converge. The vaccine counts fall just on the convergent side, so the nine intervals nearly agree on both the VE bounds and their coverage. Their differences appear only at sparser counts. A comparison with many cases in *both* arms would be moot.

The Covid-19 readouts serve only as a familiar, publicly documented instance of this sparse regime: any two-arm rare-event count comparison with few events in one arm (an early drug-safety signal, a reliability test with few failures, an interim oncology readout) would make the same points. We use these data because the counts are public and well known, not because the construction is specific to vaccines or to Covid-19.

### Moderna mRNA-1273 (COVE trial, Nov 30, 2020)

*K* = 11 cases in the vaccine arm, *L* = 185 in placebo (*N* = 30,000 participants), giving 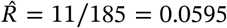 and 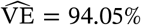. These are the primary efficacy-analysis counts reported by Fu et al.; the methodology applies identically to the earlier interim readout (5 vaccine, 90 placebo) or to any later one. We evaluate the recommended interval (19) on these counts, term by term. With *z*_0.975_ = 1.96,

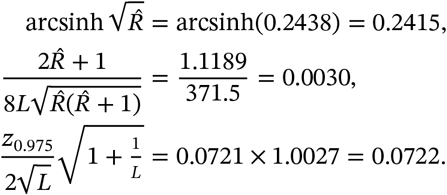

The corrected center is 0.2415 + 0.0030 = 0.2445. Subtracting and adding the half-width gives 0.1723 and 0.3167, and sinh^2^ of each returns the bounds on the *R* scale:

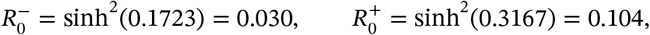

so VE = 1 − *R* ∈ (0.896, 0.970), the +Curve+Stu row of Table 6.

**TABLE 6.**
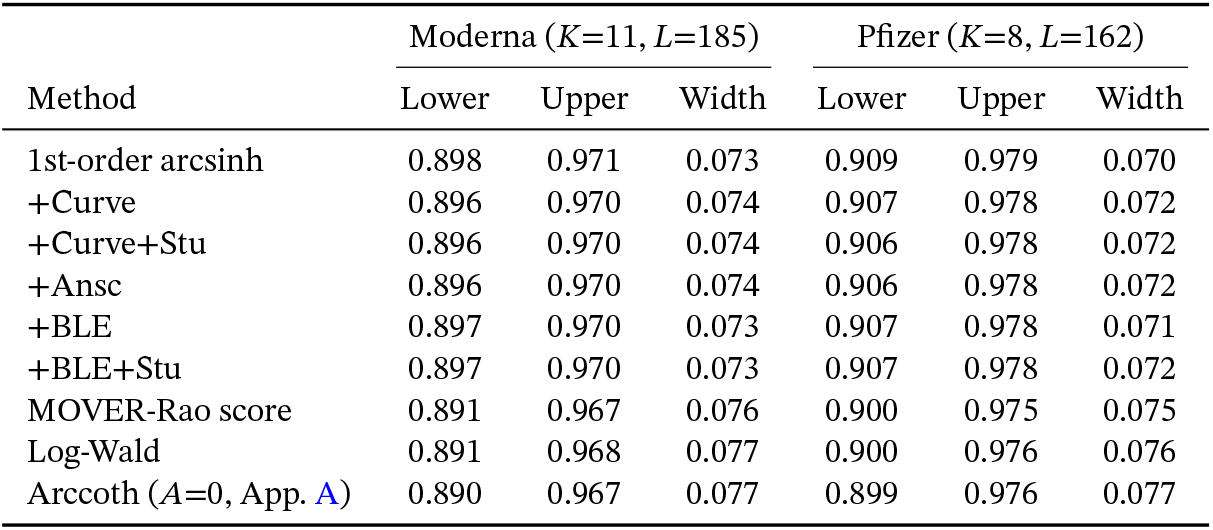
95% CI for vaccine efficacy VE = 1 − *R*: Moderna (*K* = 11, *L* = 185) and Pfizer (*K* = 8, *L* = 162).

| Method | Moderna ($K=11, L=185$) | | | Pfizer ($K=8, L=162$) | | |
| --- | --- | --- | --- | --- | --- | --- |
|  | Lower | Upper | Width | Lower | Upper | Width |
| 1st-order arcsinh | 0.898 | 0.971 | 0.073 | 0.909 | 0.979 | 0.070 |
| +Curve | 0.896 | 0.970 | 0.074 | 0.907 | 0.978 | 0.072 |
| +Curve+Stu | 0.896 | 0.970 | 0.074 | 0.906 | 0.978 | 0.072 |
| +Ansc | 0.896 | 0.970 | 0.074 | 0.906 | 0.978 | 0.072 |
| +BLE | 0.897 | 0.970 | 0.073 | 0.907 | 0.978 | 0.071 |
| +BLE+Stu | 0.897 | 0.970 | 0.073 | 0.907 | 0.978 | 0.072 |
| MOVER-Rao score | 0.891 | 0.967 | 0.076 | 0.900 | 0.975 | 0.075 |
| Log-Wald | 0.891 | 0.968 | 0.077 | 0.900 | 0.976 | 0.076 |
| Arccoth ($A=0$, App. A) | 0.890 | 0.967 | 0.077 | 0.899 | 0.976 | 0.077 |

For a trial whose arms differ in exposure, the *R*-scale endpoints multiply by the exposure ratio *τ* = *t*_0_/*t*_1_ (Section 1.3). A control arm with 5% more exposure (*τ* = 1.05) would give, for these counts, *R* ∈ (0.031, 0.109) and VE ∈ (0.891, 0.969). The readouts here have equal exposure, so *τ* = 1.

### Pfizer-BioNTech BNT162b2 (Nov 18, 2020)

*K* = 8 cases in the vaccine arm, *L* = 162 in placebo (*N* = 43,000 participants), giving 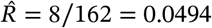 and 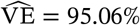.

### The original analyses

The two trials estimated efficacy by different routes, both person-time based. Pfizer used the incidence rate ratio, with a Bayesian beta-binomial credible interval and a Clopper–Pearson confidence interval ^2^. That is the conditional-binomial route, and the arcsinh interval here is its unconditional counterpart. Moderna used a Cox proportional-hazards model ^3^. The reported efficacies are essentially the point estimate 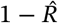. Both trials also gave 95% intervals, from the beta-binomial and the Cox fit. Table 6 gives ours in closed form.

### Regulatory criterion

The FDA’s Emergency Use Authorization (EUA) guidance for Covid-19 vaccines ^23^ required the *lower bound of the two-sided 95% confidence interval for vaccine efficacy* to exceed 30%. Since VE = 1 − *R*, this corresponds to 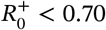 on the rate-ratio scale (where 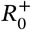 is the upper bound of the 95% CI for *R* under the standard 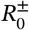 convention). All methods in Table 6 exceed this threshold for both Moderna and Pfizer. More to the point, the criterion is why a CI lower bound, not the point estimate, is the policy-relevant quantity.

For the Moderna data (*B* = 185, *BR* = 11), the +Curve+Stu CI (0.896, 0.970) is slightly narrower than MOVER-Rao (0.891, 0.967) and log-Wald (0.891, 0.968). +BLE (0.897, 0.970) is essentially indistinguishable from +Curve+Stu but with the simpler shifted-count form (*K* + 1/4)/(*L* + 1/2). The Pfizer data (*B* = 162, *BR* = 8) shows the same pattern at slightly wider intervals due to fewer events. In both cases, all methods confirm high vaccine efficacy. The *A* = 0 arccoth alternative is only marginally wider here, a second-order 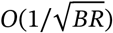 difference that grows as counts fall. Its construction and the full comparison are in Appendix A.4.

At the Moderna data point, the exact conditional coverage is: +Curve+Stu 0.949, +Ansc 0.950, +BLE 0.948, MOVER-Rao 0.952, log-Wald 0.957, all near nominal (0.948–0.952) apart from log-Wald’s mild conservatism. At the Pfizer data point (*BR* = 8, in the sparse part of the recommended *BR* ≥ 5 regime), +Curve+Stu still achieves 0.949, demonstrating robustness for moderate event counts.

## 6 Discussion

### 6.1 Summary of findings

The arcsinh VST provides a closed-form confidence interval for the Poisson rate ratio that, after second-order correction, covers within 0.002 of the nominal 0.95 for *B* ≥ 50, *BR* ≥ 5. The corrections decompose into two independently interpretable and independently derivable terms:

1. A *curvature correction* from *g*^′′^ of the transformation, which shifts the interval center by 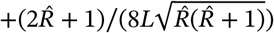.
2. A *studentization correction* 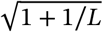 from the random studentization effect (the Poisson-*t* analogue), which inflates the half-width.

Neither correction carries a tuning constant. The curvature shift is derived from the bivariate delta expansion. The studentization inflation 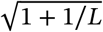 is the leading tail-widening correction of Student-*t* type, confirmed by the coverage study.

### 6.2 Connections to the literature

The half-width factor 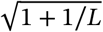 is a *studentization* correction of Student-*t* type, not a Bartlett correction: it widens the normal quantile toward its Student-*t* counterpart, treating the single count *L* as the carrier of scale information, and needs no higher-order likelihood theory. The rate-ratio deviance’s Bartlett coefficient is instead *R*-dependent (Appendix B).

The +Ansc and +BLE forms (methods (e) and (f)) achieve comparable coverage through a different mechanism : shifting the counts before the VST rather than the pivot after it. Both descend from the classical Anscombe result ^4^ that 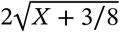 is variance-stabilized for *X* ~ Pois(*μ*). The Bar-Lev–Enis construction sharpens it by choosing the shifts (*a, b*) specifically for the rate ratio, giving the unique (1/4, 1/2) that kills the leading *O*(1/*B*) bias uniformly in *R* (derived in Section 3.3). The arcsinh CI for either 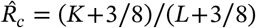 or 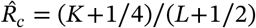 therefore needs no separate curvature correction. The count shift zeroes the leading finite-sample bias at the individual-count level rather than expanding around it. Unlike the pivot corrections, it therefore does not rely on *ε* ≪ 1. That, together with the higher one-sided bound the shifted center (*K*+*a*)/(*L*+*b*) keeps at *K* = 0 (Section 4), is why +Ansc and +BLE stay robust for *BR* < 5, where the pivot corrections break down.

#### Profile likelihood

The +Curve+Stu CI matches the profile likelihood-ratio interval’s finite-sample coverage in closed form, without the iterative root-finding that interval requires ^24^. The profile CI inverts the deviance statistic 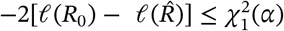 and is asymptotically equivalent to the arcsinh pivot to first order. The second-order corrections carry that agreement into finite samples, all in closed form.

#### Asymptotic efficiency

Since the pivot (8) is constructed from the MLE 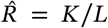 via the VST 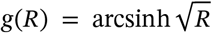 satisfying 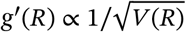, the resulting CI is asymptotically efficient: its expected width scales as 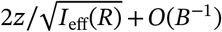, matching the Cramér–Rao bound to leading order.

#### Connection to the binomial CI

Conditioning on *N* = *K*+*L* yields *K* ∣ *N* ~ Bin(*N, p*) with *p* = *R*/(*R*+1), a one-parameter model free of *B*. A normal-approximation CI uses the binomial VST arcsin 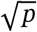 with variance ≈ 1/(4*N*):

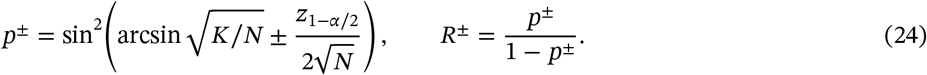

Our unconditional arcsinh CI (9) uses the Poisson ratio VST arcsinh 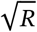 instead. The two are related by the identity

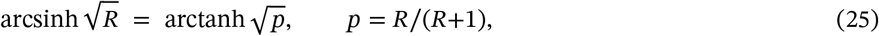

which differs from the binomial 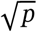 by a third-order term: both equal 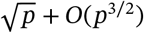 for small *p*. Correspondingly, the arcsinh CI uses sinh^2^ while the binomial CI uses sin^2^ / cos^2^ = tan^2^ for the back-transformation. For rare events (*p* ≪ 1, *N* ≈ *L*), we have sin ≈ sinh ≈ tan, and the two CIs converge:

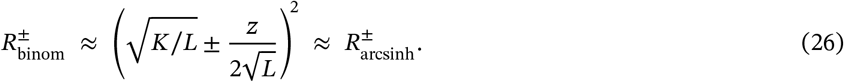

Thus, where the textbook route conditions on *N* = *K*+*L* and falls back on the binomial arcsine CI, the construction here is *unconditional* (it stabilizes the rate ratio directly, on the arcsinh 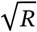 scale). The two coincide only in the rare-event limit typical of vaccine efficacy studies. The +Ansc continuity correction (*K* + 3/8, *L* + 3/8) is a continuity-corrected approximation to the binomial interval. The +BLE shift (*K*+1/4, *L*+1/2) is the bias-killing variant tailored to the bivariate ratio rather than the single Poisson.

### 6.3 Overdispersion and the negative-binomial neighbor

The effective single-parameter problem’s immediate neighbor is the negative binomial. The two share a variance function. The negative binomial is the NEF-QVF member ^7^ whose variance exceeds its mean. Under extra-Poisson variation, it describes the counts better than the Poisson does. The same two-step refinement then applies with a quasi-Poisson dispersion factor or the negative-binomial variance in place of the Poisson one.

Concretely, take the quasi-Poisson case. The variance picks up a dispersion factor *ϕ*,

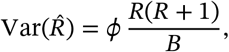

so every standard error on the stabilized scale scales by 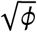. The half-width of Eq. (19) becomes 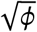 times its Poisson value. The curvature correction (i.e. +Curve), linear in the plugged-in variance, picks up the same factor *ϕ*. What this needs is an estimate 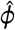, and one readout cannot supply it. A single count per arm does not identify *ϕ*. Estimating it takes residual degrees of freedom, from a Poisson or negative-binomial regression across strata, not the two counts of one readout. More generally, the curvature correction is the generic delta-method term 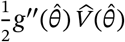 for the VST kernel 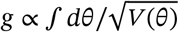 and carries over to any NEF-QVF variance-stabilizer. The studentization ^2^(+Stu), by contrast, depends on the joint law of the estimator and its scale statistic and must be re-derived per model.

### 6.4 Limitations

Four limits mark where the guarantees end:

1. All coverage guarantees are conditional on the Poisson model. Under overdispersion, the intervals run narrow (Section 6.3). A single count per arm does not identify the dispersion, so correcting it needs an external scale estimate, not an internal one.
2. The curvature correction breaks down when *BR* ≲ 5. The Taylor expansion needs enough events to be accurate. Conditional coverage there is non-monotone. It troughs to about 0.70 near *BR* ≈ 1.25, then rebounds above nominal at smaller *BR*. The rebound is an artifact of the *K* = 0 boundary, not reliable coverage (Key Observations, Section 4.5). This is the boundary of the interval’s validity, and the reason the count-shift forms (+BLE preferred, Section 3.3; +Ansc; MOVER-Rao) take over below *BR* = 5.
3. The studentization correction slightly overshoots for *B* ≤ 10 (coverage ~ 0.96, conservative and hence safe), because 1 + 1/*L* overestimates the true tail-widening effect for very small *B*.
4. The CI is undefined when *L* = 0 (no control events). This is inherent to the unconditional formulation.

*Remark 2. One-sided intervals*. One-sided CIs are obtained by replacing *z*_1−*α*/2_ with *z*_1−*α*_ in Eq. (19) and retaining only the relevant bound. All coverage properties transfer directly.

## 7 Conclusion

The bivariate Poisson rate ratio lies outside the single-variate NEF-QVF classification. The *A* = 1 reparameterization brings it within reach: it renders the control arm free of *R* and recasts the problem in an effective single-parameter, quadratic-variance form, with *A* = 1 the preferred exponent at rare events (Appendix A). On that effective problem we build the arcsinh interval and two closed-form corrections with no tuning constants: a curvature-bias shift, derived from the delta expansion, and a studentization inflation of Student-*t* type, confirmed by the coverage study. The corrected interval (Eq. 19) attains near-nominal coverage in the moderate-count regime.

For routine use we recommend the +Curve+Stu interval where *B* ≥ 50 and *BR* ≥ 5, and a count-shift form in the sparse regime (Section 4.7). The curvature correction carries over to any NEF-QVF variance-stabilizer. The studentization must be re-derived per model.

## Data Availability

All code and simulated data produced in this study are openly available. Python
and R code for all confidence-interval methods, Monte Carlo simulations, and
figure and table generation is available at
https://github.com/ovect/arcsinh-poisson-rate-ratio and archived at Zenodo
(https://doi.org/10.5281/zenodo.21331365). An independent R reimplementation
reproduces the main coverage table to within a few thousandths (maximum
deviation 0.001 across all entries). The Covid-19 vaccine trial data are two
published aggregate event counts from the primary trial reports (Baden et al.
2021; Polack et al. 2020).

https://github.com/ovect/arcsinh-poisson-rate-ratio

https://doi.org/10.5281/zenodo.21331365

https://doi.org/10.1056/NEJMoa2035389

https://doi.org/10.1056/NEJMoa2034577

## Acknowledgments

This research was self-funded and conducted independently. The author declares no competing interests.

## Data Availability Statement

Python and R code for all CI methods, Monte Carlo simulations, and figure and table generation is available at https://github.com/ovect/arcsinh-poisson-rate-ratio and archived at Zenodo (https://doi.org/10.5281/zenodo.21331365). An independent R reimplementation reproduces Table 3 to within a few thousandths (maximum deviation 0.001 across all entries). The Covid-19 vaccine trial data are from the primary trial reports ^3,2^.

## APPENDIX A A The *A*-Parameterization and the Choice of *A* = 1

### A.1 General framework

To justify *A* = 1 we work the whole *A*-line and compare. Under the parameterization (4) with arbitrary *A*, the delta-method variance of 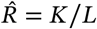 is

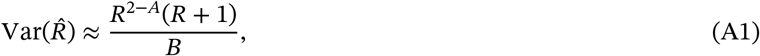

and the VST integrand becomes 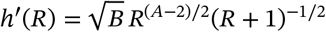.

The antiderivative is elementary for every integer *A* ^25^. But only the four “base” exponents *A* ∈ {−1, 0, 1, 2} yield a transformation with a *single-term* elementary inverse. The confidence interval requires that inverse, since the bounds are obtained by inverting the pivot:

| $A$ | VST $h(R)/\sqrt{B}$ | CI inverse | $R$-free $\hat{B}$? |
| --- | --- | --- | --- |
| -1 | $-2\sqrt{1+1/R}$ | $1/[(\dots)^2 - 1]$ | No ($\hat{B} = K^2/L$) |
| 0 | $-2 \operatorname{arccoth} \sqrt{R+1}$ | $1/\sinh^2(\dots)$ | Yes ($\hat{B} = K$) |
| 1 | $2 \operatorname{arcsinh} \sqrt{R}$ | $\sinh^2(\dots)$ | Yes ($\hat{B} = L$) |
| 2 | $2\sqrt{R+1}$ | $(\dots)^2 - 1$ | No ($\hat{B} = L^2/K$) |

These four are exactly the exponents for which the substitution 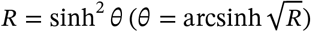 reduces the VST integral to a *single* elementary term, 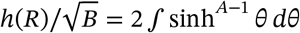, with the power-reduction recursion terminating in one step. The four base cases come in two dual pairs under the arm-swap *R* ↔ 1/*R* (equivalently *A* ↔ 1 − *A*, which interchanges *K* and *L*), related by *h*_1−*A*_(1/*R*) = −*h*_*A*_(*R*). The pairs are the inverse-hyperbolic {*A* = 1, *A* = 0} 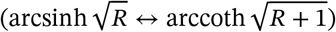 and the algebraic 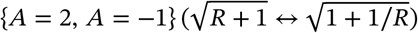. The two exponents in a pair are the same transformation read from opposite arms. The choice of *A* = 1 over its partners rests on the clean, *R*-free nuisance and effective sample size (next subsection), not on the closed form, which all four share. The same sinh-power reduction yields the (non-invertible) closed form at every other integer *A*.

### A.2 Why *A* = 1 is preferred

Three properties recommend *A* = 1:

1. **Parameter orthogonality and a clean nuisance**. With the nuisance taken to be the total intensity *λ* = *B*(*R* + 1) = *E*[*K* + *L*], the parameters *R* and *λ* are exactly orthogonal in the sense of Cox and Reid ^26^ (the Fisher information is diagonal; Section A.3). At *A* = 1 the control arm *L* ~ Pois(*B*) has a distribution free of *R* (*E*[*L*] = *BR*^*A*−1^ = *B* is *R*-free only at *A* = 1), so 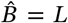 carries no information about *R*. The dual exponent *A* = 0 shares this clean single-channel structure under the arm swap *R* ↔ 1/*R*: there 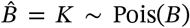 is the *R*-free arm. For any other *A*, both arms involve *R*, and the nuisance estimator 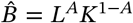 mixes the two channels. *A* = 1 is fixed over its dual by the effective-sample-size property below.
2. **Largest effective sample size**. The studentization uses 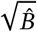, and for rare events (*R* ≪ 1) the control arm is the larger count. At *A* = 1 the nuisance estimator is that larger arm, 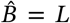 at *A* = 0 it is the smaller treatment arm, 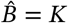. The vaccine example (*K* = 11, *L* = 185) studentizes by 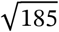 at *A* = 1 versus 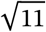 at *A* = 0.
3. **Conditional sufficiency**. Conditioning on *N* = *K* + *L* yields *K* ∣ *N* ~ Bin(*N, R*/(*R* + 1)), which is entirely *B*-free: the nuisance parameter has been eliminated by conditioning, reducing the problem to a standard one-parameter binomial model. This provides both an exact (conditional) CI via the binomial and the theoretical guarantee that no information about *R* is lost in the studentization step. Unlike the first two properties, this conditional reduction is not special to *A* = 1: it depends only on the ratio of arm means *μ*_1_/*μ*_0_ = *R* and so holds for every *A*. It is listed here because it underwrites the studentization and supplies the exact conditional reference of Section 4. The choice of *A* = 1 itself is fixed by the clean-nuisance and effective-sample-size properties above.

### A.3 Parameter orthogonality and the Fisher information

Take the nuisance to be the total intensity *λ* = *B*(*R* + 1) = *E*[*K* + *L*] (treatment plus control intensity), estimated by *N* = *K* + *L*, and write *p* = *R*/(*R* + 1). Then *K* ~ Pois(*λp*) and *L* ~ Pois(*λ*(1 − *p*)), and the log-likelihood separates,

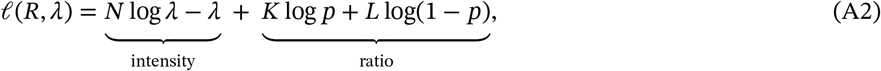

so the Fisher information for (*R, λ*) is exactly diagonal,

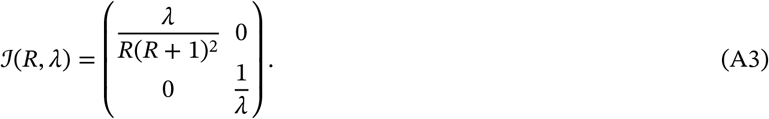

Its diagonal entry ℐ_*RR*_ = *λ*/[*R*(*R* + 1)^2^] = *B*/[*R*(*R* + 1)] is the effective information for *R*, from which the variance-stabilizing rule 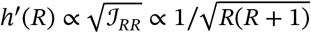 recovers the transform (6) directly. Because the off-diagonal vanishes, *R* and *λ* are orthogonal parameters in the sense of Cox and Reid ^26^. An orthogonal nuisance always exists for a scalar parameter of interest, and here it is the clean, interpretable *λ* = *E*[*N*].

The vanishing off-diagonal does *not* make 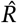 more precise. The marginal asymptotic variance is invariant to the parameterization, 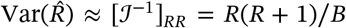 (Eq. (5)), the same value the (*R, B*) information gives. What orthogonality buys is that *λ* is a *cost-free* nuisance: knowing the total intensity would not sharpen 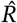 at all, since all information about *R* sits in the *λ*-free conditional split *K* ∣ *N* ~ Bin(*N, p*). Contrast *B*, an *informative* nuisance: were *B* known, 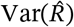 would fall from *R*(*R* + 1)/*B* to *R*/*B*, a factor of *R* + 1.

### A.4 The *A* = 0 alternative: simulation comparison

The *A* = 0 CI uses 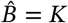 and inverts the arccoth VST:

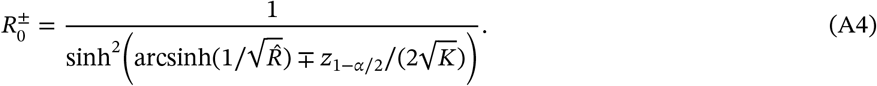

Table A1 compares *A* = 0 and *A* = 1 coverage.

The first-order *A* = 0 CI achieves near-nominal coverage *when it is defined*, without second-order corrections. Its studentizer is the treatment-arm count 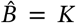, and the 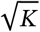 scale needs no curvature or studentization fix as long as *K* stays clear of zero. The *K* = 0 boundary is its only weakness. On the common data-generating process, the treatment mean is *E*[*K*] = *BR*, so the arccoth interval is undefined with probability *P*(*K* = 0) = *e*^−*BR*^ (last column of Table A1). That probability is negligible when *BR* is large, but reaches 8% at *B* = 25, *R* = 0.1 and 5% at *B* = 50, *R* = 0.06. The strong entries in Table A1 are therefore *conditional* coverages: the arccoth CI trades the always-defined guarantee of the arcsinh interval for a boundary hole that widens as *BR* → 0. For *R* ≤ 1 known a priori (e.g. vaccine efficacy) with a treatment arm that is not too sparse (as at the Moderna point, *K* = 11), the arccoth CI is thus a strong, correction-free alternative to +Curve+Stu. Only when *BR* ≲ 3 does its boundary hole make the always-defined arcsinh interval the safer choice. For *R* > 1, second-order corrections would in principle apply (by the arm swap *A* ↔ 1 − *A* they mirror the *A* = 1 corrections), but we do not pursue them here.

**TABLE A1.** Coverage comparison under the single *A* = 1 data-generating process (*K* ~ Pois(*BR*), *L* ~ Pois(*B*)), so all three CIs are evaluated on the *same* data: the *A* = 0 arccoth CI, the first-order *A* = 1 arcsinh CI, and log-Wald. Coverage is conditional on the interval being defined (as in Table 3). The last column is the arccoth CI’s boundary rate on this data, *P*(*K*=0) = *e*^−*BR*^, the fraction of samples on which arccoth is undefined. Exact Poisson summation (Section 4).

| $B$ | $R$ | Arccoth CI ($A=0$) | Arcsinh CI ($A=1$) | Log-Wald | $P(K=0) = e^{-BR}$ |
| --- | --- | --- | --- | --- | --- |
| 10 | 0.50 | 0.955 | 0.938 | 0.972 | $6.7 \times 10^{-3}$ |
| 10 | 2.00 | 0.945 | 0.954 | 0.958 | $2.1 \times 10^{-9}$ |
| 25 | 0.10 | 0.958 | 0.906 | 0.964 | $8.2 \times 10^{-2}$ |
| 25 | 1.00 | 0.951 | 0.951 | 0.954 | $1.4 \times 10^{-11}$ |
| 50 | 0.06 | 0.962 | 0.934 | 0.964 | $5.0 \times 10^{-2}$ |
| 100 | 0.06 | 0.965 | 0.939 | 0.967 | $2.5 \times 10^{-3}$ |
| 185 | 0.06 | 0.956 | 0.945 | 0.956 | $1.5 \times 10^{-5}$ |
| 500 | 0.05 | 0.952 | 0.948 | 0.953 | $1.4 \times 10^{-11}$ |
| 500 | 1.00 | 0.950 | 0.950 | 0.950 | $7 \times 10^{-218}$ |

#### Coupling at *A* = 0

The arccoth CI’s nuisance estimator is 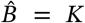, which the pivot 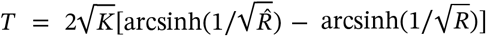 uses twice: through 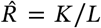 and through the studentizing factor 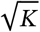. So 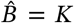 and 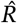 are correlated. This coupling is not peculiar to *A* = 0: at *A* = 1 the estimators 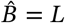 and 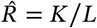 are likewise correlated through their shared *L*, and the Fisher information for (*R, B*) is non-diagonal at both exponents. Exact orthogonality holds only in the (*R, λ*) reparameterization of Section A.3. What differs between the two is which arm does the studentizing. How reliable that arm is for *R* ≪ 1 is the subject of the next paragraph.

For rare events (*R* ≪ 1) the treatment arm is the *smaller* count, so 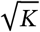 is a noisier scale than the 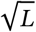 of *A* = 1 and degrades as *K* → 0, where the arccoth interval is undefined. The VE intervals are comparable at the Moderna data (*K* = 11, *L* = 185): (0.890, 0.967) at *A* = 0 versus (0.898, 0.971) at *A* = 1, because the steeper arccoth back-transform offsets *A* = 0’s wider studentized half-width. The *A* = 1 advantage is thus one of studentizer reliability, growing as *B* falls and *K* nears its boundary.

## B The Bartlett-corrected likelihood-ratio interval

The natural second-order competitor to the closed-form VST interval is the Bartlett-corrected likelihood-ratio (LR) interval. We give its construction and note that its Bartlett coefficient is *R*-dependent (so it is *not* the 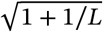 factor of Section 3).

### Deviance

Under *K* ~ Pois(*BR*), *L* ~ Pois(*B*) the log-likelihood is *ℓ*(*R, B*) = (*K* + *L*) log *B* + *K* log *R* − *B*(*R* + 1), with unconstrained 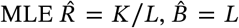 and profile scale 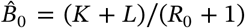 under *H*_0_ : *R* = *R*_0_. The deviance is the Poisson *G*^2^,

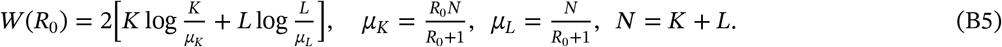

The Poisson likelihood factors into an *N*-marginal and a binomial term, and at 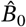 the marginal is fitted exactly. Hence *W*(*R*_0_) equals the binomial deviance for *K* ∣ *N* ~ Bin(*N, p*_0_), *p*_0_ = *R*_0_/(*R*_0_ + 1). The LR interval is 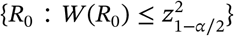, found by root-finding on each side of 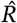 (no closed form), then mapping *p*_0_ → *R*_0_ = *p*_0_/(1 − *p*_0_).

### Bartlett coefficient

A fourth-order expansion of the binomial deviance gives *E*[*W* ∣ *N*] = 1 + *b*(*p*_0_)/*N* + *O*(*N*^−2^) with *b*(*p*) = (1 − *p* + *p*^2^)/[6*p*(1 − *p*)]. Averaging over *N* ~ Pois(*B*(*R* + 1)) yields

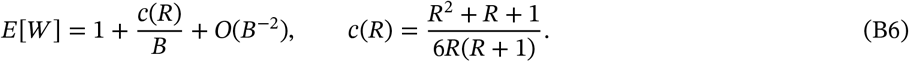

The Bartlett-corrected interval replaces the cutoff 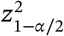 by 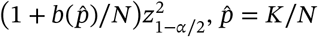.

### Not the studentization factor

The coefficient *c*(*R*) is *R*-dependent, running from *c*(1) = 1/4 to *c*(∞) = 1/6 for *R* ≥ 1 (0.17–0.25) and growing like 1/(6*R*) as *R* → 0. It is never the unit value the 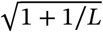 studentization factor of Section 3 would need to be a Bartlett correction. The two act on different statistics (a Bartlett adjustment of the LR deviance versus a Student-*t* tail correction of the VST pivot), coinciding only in being *O*(1/*B*) refinements.

## Footnotes

1 We write *B* for the scale rather than *n*: the operative quantity is the Poisson exposure (*E*[*L*] = *B* at *A* = 1), and the large-sample limit is *B* ---+ ∞, not a count of independent replicates.

2 Moments of 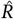 are taken conditional on *L* ≥ 1 throughout: unconditioned, *E*[1/*L*] and hence 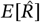 diverge at *L* = 0. The conditioning event has probability *e*^−*B*^ and shifts every displayed coefficient by *O*(*e*^−*B*^) only, far below the *O*(*B*^−2^) truncation.

